# Sex-specific associations of childhood socioeconomic position and trajectories of metabolic traits across early life: prospective cohort study

**DOI:** 10.1101/2022.05.09.22274827

**Authors:** Kate N O’Neill, Joshua A Bell, George Davey Smith, Abigail Fraser, Laura D Howe, Patricia M Kearney, Oliver Robinson, Kate Tilling, Peter Willeit, Linda M O’Keeffe

## Abstract

**Background:** Socioeconomic inequalities in cardiovascular disease risk begin early in life and are more pronounced in females compared with males in later life but the causal atherogenic traits that may explain this are not well understood. We explored sex-specific associations between indicators of childhood socioeconomic position (SEP) and changes in molecular measures of systemic metabolism across early life.

**Methods:** Data were from offspring of the Avon Longitudinal Study of Parents and Children (ALSPAC), born in 1991/1992. Maternal education was the primary indicator of SEP with paternal education and household social class used as secondary indicators; all measures were collected through questionnaires administered to mothers at 32-weeks’ gestation of the offspring pregnancy. Concentrations of 148 metabolic traits were measured using nuclear magnetic resonance spectroscopy performed on plasma samples at ages 7 years (y), 15y, 18y and 25y among offspring. The sex-specific slope index of inequality (SII) in trajectories of metabolic traits across these ages was estimated using multilevel models.

**Results:** Between 6,010-6,537 participants with 10,055-12,543 repeated measures of metabolic traits from 7y to 25y were included. Lower maternal education was associated with more adverse levels of several atherogenic lipids and other key metabolic traits among females at age 7y, but not males. For instance, the SII for very small very-low-density lipoprotein (VLDL) concentrations was 0.16 SD (95% CI: 0.01, 0.30) among females and -0.02 SD (95% CI: -0.16, 0.13) among males at 7y. Between 7y and 25y, inequalities widened among females and emerged among males particularly for VLDL particle concentrations, plasma apolipoprotein B concentrations and inflammatory glycoprotein acetyls. For instance, at 25y the SII for very small VLDL concentrations was 0.36 SD (95% CI: 0.20, 0.52) and 0.22 SD (95% CI: 0.04, 0.40) among females and males respectively. Findings for secondary SEP indicators were broadly similar although associations of paternal education with key metabolic traits were weak and less consistent among males at 25y compared with associations of maternal education.

**Conclusion:** Socioeconomic inequalities in causal atherogenic lipids and other key metabolic traits such as markers of inflammation begin in childhood and strengthen in adolescence among females but only emerge in adolescence among males, leading to wider socioeconomic inequalities among females compared with males by 25y. Prevention of socioeconomic inequalities in cardiovascular disease risk requires a life course approach that begins at the earliest opportunity in the life course especially among females.

## Introduction

Socioeconomic position (SEP) captures multiple dimensions of social inequality usually defined by educational attainment, occupational class and/or material circumstances (1,2). Socioeconomic inequalities in cardiovascular disease (CVD) risk are well established (3–5) with adults of disadvantaged SEP experiencing a higher incidence of CVD morbidity and mortality (3,6). Inequalities in CVD risk likely manifest early in the life course (7–11), and appear to be stronger and more consistent in females compared with males (3,12). For example, a systematic review and meta-analysis of prospective cohort studies with over 22 million individuals found that disadvantaged SEP in females carries a 34% excess risk of coronary heart disease incidence compared with disadvantaged SEP in males (12).

Recent studies have focussed on identifying modifiable risk factors for CVD such as adiposity (often measured using body mass index [BMI]) that may mediate the effect of SEP on CVD risk across the life course (13,14). However, while three established risk factors – adiposity, smoking, and systolic blood pressure – may explain up to half of the effect of SEP on CVD (14), the mechanisms mediating the remaining effect of SEP are still unknown (3,14,15). Advances in targeted metabolomics methods and their application to population-based cohort studies present new opportunities to understand the more fine-grained molecular pathways that may mediate SEP-CVD risk (16). A recent multi-cohort analysis of approximately 30,000 adults and 4,000 children in the UK and Finland reported associations between SEP and a number of molecular traits that are known to cause and/or predict CVD (9,17,18). For instance, associations of father’s occupation with molecular metabolic traits such as omega-3 fatty acids and free cholesterol as a percentage of very large high-density lipoprotein (HDL) were already evident at age 7 years, and were similar to socioeconomic inequalities previously observed for adiposity (7,19). However, other dimensions of childhood SEP such as maternal education were not explored in this study and associations were not reported separately in females and males, despite evidence of stronger associations between disadvantaged SEP and higher CVD risk in females (7,12). In addition, associations between SEP and trajectories of these traits throughout childhood and adolescence into adulthood were not examined. Examining trajectories can determine when in the life course SEP begins to influence CVD susceptibility and how the influence of SEP on CVD risk may change over time. This understanding can provide insights into the aetiology and potential mediating pathways, e.g., differences that are evident in early childhood may implicate familial factors related to early life adversity while differences emerging around puberty could suggest a role for adiposity or health behaviours of the child.

Using data from a prospective birth cohort study in the southwest of England, we examined sex-specific associations between early-life indicators of SEP and trajectories of 148 molecular measures of systemic metabolism (mostly concentrations of lipoprotein subclasses, fatty acids, amino acids, fluid balance factors, glycolysis-related metabolites including glucose, ketone bodies, and inflammatory glycoprotein acetyls (GlycA)) measured using targeted metabolomics on four occasions among the same individuals, from ages 7 to 25 years (y).

## Methods

### Study population

Data were from first-generation children of the Avon Longitudinal Study of Parents and Children (ALSPAC), a population-based prospective birth cohort study in southwest England. Pregnant women resident in one of the three Bristol-based health districts with an expected delivery date between April 1, 1991, and December 31, 1992, were invited to participate. ALSPAC initially enrolled a cohort of 14,451 pregnancies, from which 14,062 live births occurred, and 13,988 children were alive at 1 year of age. When the oldest children were approximately 7 years of age, an attempt was made to bolster the initial sample with eligible cases who had not joined the study originally. Therefore, the total sample size for analyses using any data collected after the age of 7 is 14,901 children. Follow-up has included parent- and child-completed questionnaires, research clinic attendance, and links to routine data. Data gathered from participants at 22 years of age and onwards were collected and managed using REDCap (Research Electronic Data Capture) electronic data capture tools (20,21). Ethical approval for the study was obtained from the ALSPAC Ethics and Law Committee and the Local Research Ethics Committees. Informed consent for the use of data collected via questionnaires and clinics was obtained from participants following the recommendations of the ALSPAC Ethics and Law Committee at the time. The study website contains details of all the data that is available through a fully searchable data dictionary http://www.bristol.ac.uk/alspac/researchers/our-data/.

### Socioeconomic position

#### Primary indicator

Maternal educational attainment was used as the primary indicator of SEP as it is the most complete measure available and is the most frequently reported indicator of childhood SEP (7,10,22). At 32-weeks’ gestation, mothers were asked to report their highest educational attainment based on UK standards at the time which was categorised as ‘less than O-level’ (Ordinary Level; exams taken in different subjects usually at age 15y or 16y at the completion of legally required school attendance, equivalent to the present UK General Certificate of Secondary Education; assumed to reflect the lowest SEP), ‘O-level’, ‘A-level’ (Advanced Level; exams taken in different subjects usually at age 18 years), or ‘university degree’ (undergraduate or postgraduate; assumed to reflect the highest SEP).

#### Secondary indicators

We also explored two other dimensions of childhood SEP: mothers’ partner’s educational attainment (paternal education) and household social class. Mothers were asked to report their partner’s educational attainment at 32-weeks’ gestation which was categorised in the same way as maternal education. Household social class was measured as the highest of the mother’s or her partner’s occupational social class using data on job title and details of occupation collected about the mother and her partner from the mother’s questionnaire at 32-weeks’ gestation. Occupation was categorised into four categories for this analysis according to the standard occupational classification (SOC) codes developed by the United Kingdom Office of Population Census and Surveys: ‘professional’ (assumed to reflect the highest SEP), ‘managerial and technical’, ‘non-manual’, or ‘manual, part skilled occupations and unskilled occupations’ (assumed to reflect the lowest SEP).

### Assessment of metabolic traits

Targeted metabolomics, using a proton nuclear magnetic resonance (^1^H-NMR) spectroscopy based platform (23), was performed on EDTA-plasma samples from blood samples drawn in clinics at ages 7y, 15y, 18y and 25y to quantify concentrations of 148 molecular traits, including lipoprotein subclasses, fatty acids, amino acids, fluid balance factors including glucose, glycolysis-related metabolites, ketone bodies, and inflammatory GlycA. Four trait concentrations (diacylglycerol, fatty acid chain length, estimated degree of unsaturation, and conjugated linoleic acid) were not measured at 25y; thus changes in these traits over time are only modelled to age 18y. Bloods were taken after a minimum of a 6-hour fast, except for the 7y occasion which was non-fasting (stability in these trait concentrations has been shown over different fasting durations (24)). Laboratory ^1^H-NMR quality control and further data preparation steps are described in Supplementary Material eMethods 1.

### Participants eligible for analysis

Participants with data on sex, maternal education and at least one measurement of the 148 metabolic concentrations between 7y and 25y were included in the primary analysis. Measures taken on participants who reported being pregnant at the 18-year clinic (N=3) or 25-year clinic (N=9) were recoded as missing at that timepoint.

### Statistical analysis

We used multilevel models to examine the sex-specific association between SEP and trajectories of 144 metabolic trait concentrations from 7y to 25y and four metabolic trait concentrations from 7y to 18y only (diacylglycerol, fatty acid chain length, estimated degree of unsaturation, and conjugated linoleic acid) (25,26). Multilevel models estimate mean trajectories of the outcome while accounting for the non-independence (i.e. clustering) of repeated measurements within individuals and differences in the number and timing of measurements between individuals (using all available data from all eligible participants under a Missing at Random (MAR) assumption) (27,28).

Trajectories of all outcomes have been modelled previously using linear spline and linear multilevel models (two levels: measurement occasion and individual) and are summarised in Supplementary Material eMethods 2 and described elsewhere in detail (26). Linear splines allow knot points to be fit at different ages to derive periods in which change is approximately linear. In brief, the optimal model for each trait concentration was selected by comparing model fit statistics of different models and examining observed data for each concentration; these included models that assumed linear change over time to models with knot points at 15y and 18y with knot points were placed at whole years closest to mean age at clinics due to a greater density of measures. All models included interaction terms of sex with the intercept and linear splines to allow trajectories to differ in females and males, as described previously (26).

#### Sex-specific association between SEP and metabolic trajectories

To explore the sex-specific associations between maternal education and trajectories of metabolic traits, the slope index of inequality (SII) was estimated which treats SEP as a continuous measure and assumes that there are no substantial deviations from linear associations (29). We explored linearity of associations of SEP with metabolic traits by comparing regression models with maternal education as a continuous exposure to models with maternal education as a categorical exposure (using dummy variables) at each time point using a likelihood ratio test. Overall, there was little statistical evidence of departure from linearity for most traits suggesting it was appropriate to treat SEP as a continuous exposure in our analyses (eTable 2).

To estimate the SII, a rank score of maternal education from 0 to 1 was created whereby the score for those in each category is the mid-point of the cumulative proportion of the participants in that category or lower. A three-way interaction term between sex, rank score of maternal education and the intercept and each linear spline period was included in all models. This allowed the sex-specific associations of maternal education with metabolic trajectories to be estimated from a single sex-combined model by using a linear combination of main effects and interaction effects for sex, rank score of maternal education, intercepts, and slopes. The coefficient for the maternal education rank score represents the SII and can be interpreted as the mean difference in outcome between those with the lowest SEP (less than O-level maternal education) compared with the highest SEP (degree level maternal education) on the hypothetical underlying continuous distribution of maternal education. For each sex, these models directly estimate the SII for each trait level at 7y and for slopes (rate of change) from either 7y to 25y for linear models or from 7y to 15y/7y to 18y and from 15y to 25y/18y to 25y for linear spline models in original units (mostly mmol/l). Post-analysis, these estimates were used to calculate the SII in change in each trait level from 7y to 25y and the SII in each trait level at 25y. We then converted estimates to standard deviation (SD) units by dividing by the sex-combined SD of the observed metabolite in those with degree level maternal education at the respective age, to aid comparison of results between metabolites. Additional information on the models is provided in Supplementary Material eMethods 2.

Main results are presented in SD units while results in original units are presented in Supplementary Material. All trajectories were modelled in MLwiN version 3.04 (30), called from Stata version 15 using the runmlwin command (31). Data visualisation was performed using R (version 3.6.4) using the ggforestplot (0.0.2) package.

### Additional and sensitivity analyses

Analyses were repeated for our secondary indicators of SEP, paternal education and household social class. We also repeated analyses with maternal education as a categorical variable (treated as three dummy variables with university degree or above as the reference category) in the multilevel models to further examine the patterns of socioeconomic inequality across levels of SEP. We also performed weighted sensitivity analyses using inverse probability weighting to address potential selection bias (32). The participant level weights were estimated using logistic regression and were subsequently incorporated into the multilevel models as level two weights which adjust for the unequal probability of selection of the participants. The logistic regression models included sex, maternal education, household social class, maternal marital status, parity, smoking in pregnancy, birthweight, gestational age, and maternal age. Further details are included in Supplementary Material eMethods 3.

## Results

Analyses of 144 concentrations included 6,537 participants (12,543 total repeated measures; 4,605 at 7y, 2,608 at 15y, 2,640 at 18y and 2,600 at 25y) (Figure 1). There were 6,292 participants (10,465 total repeated measures; 4,815 at 7y, 2,883 at 15y and 2,767 at 18y) included in analyses of diacylglycerol, fatty acid chain length, estimated degree of unsaturation and conjugated linoleic acid as these traits were only available up to 18y. Characteristics of participants included in analyses are presented in Table 1. The distribution of maternal education was similar in females and males, with approximately 17% of mothers having attained a university degree. Compared with mothers of participants who were excluded due to missing exposure or outcome data, mothers of participants who were included in analyses were more likely to be married, have higher household social class, higher education, lower prevalence of smoking during pregnancy, lower parity, and higher maternal age (eTable 3). The number of repeat measurements available over the four occasions also varied by sex and maternal education with higher levels of loss-to-follow up observed among males and those with less than O-level maternal education (eTable 4).

**Figure 1:**
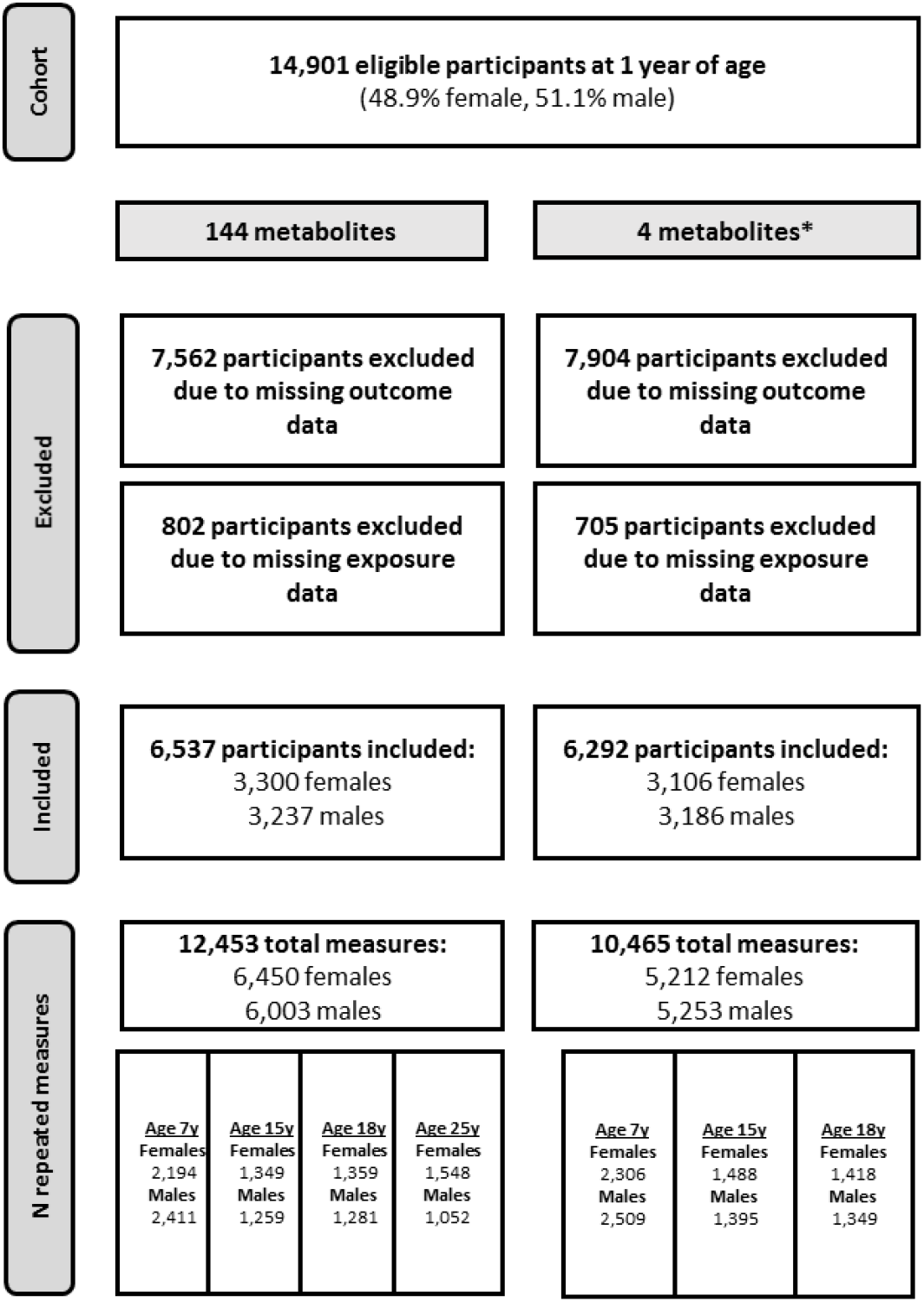
Flowchart of participants included in primary analyses. *Four metabolites (diacylglycerol, fatty acid chain length, estimated degree of unsaturation and conjugated linoleic acid) were not measured at 25y; thus, for these traits change over time is only modelled to age 18y. See Supplementary eFigures 1-2 for participants included in secondary analyses of paternal education and household social class

**Table 1.** Characteristics of ALSPAC participants included in the analysis, by sex.

|  | **Females**<br>**N=3,300** | **Males**<br>**N=3,237** |
| --- | --- | --- |
|  | **n (%)** | **n (%)** |
| **Maternal marital status\*** |  |  |
| Never married | 464(14.1) | 419(12.9) |
| Separated/Divorced/Widowed | 147(4.5) | 148(4.6) |
| 1<sup>st</sup> Marriage | 2442(74.0) | 2407(74.4) |
| Marriage 2 or more | 206(6.2) | 219(6.8) |
| Missing | 41(1.2) | 44(1.4) |
| **Household social class\*** |  |  |
| Professional | 502(15.2) | 519(16.0) |
| Managerial & Technical | 1409(42.7) | 1356(41.9) |
| Non-Manual | 765(23.2) | 765(23.6) |
| Manual, Part Skilled & Unskilled | 478(14.5) | 437(13.5) |
| Missing | 146(4.4) | 160(4.9) |
| **Maternal education\*** |  |  |
| Less than O level | 725(22.0) | 692(21.4) |
| O level | 1125(34.1) | 1158(35.8) |
| A level | 890(27.0) | 855(26.4) |
| Degree | 560(17.0) | 532(16.4) |
| **Mother's Partner's highest educational qualification\*** |  |  |
| Less than O level | 917(27.8) | 825(25.5) |
| O level | 690(20.9) | 697(21.5) |
| A level | 884(26.8) | 889(27.5) |
| Degree | 718(21.8) | 734(22.7) |
| Missing | 91(2.8) | 92(2.8) |
| **Maternal smoking during pregnancy\*** |  |  |
| Yes | 603(18.3) | 609(18.8) |
| No | 2189(66.3) | 2135(66.0) |
| Missing | 508(15.4) | 493(15.2) |
| **Parity\*** |  |  |
| 0 | 1455(44.1) | 1432(44.2) |
| 1 | 1191(36.1) | 1129(34.9) |
| 2+ | 600(18.2) | 641(19.8) |
| Missing | 54(1.6) | 35(1.1) |
| **Breastfeeding\*** |  |  |
| Exclusive | 1121(34.0) | 983(30.4) |
| Non-exclusive | 1389(42.1) | 1509(46.6) |
| Never | 510(15.5) | 496(15.3) |
| Missing | 280(8.5) | 249(7.7) |
|  | **Mean (SD)** | **Mean (SD)** |
| **Offspring gestational age (weeks)** | 39.6(1.7) | 39.3(1.9) |
| **Offspring birth weight (g)** | 3375(504.0) | 3478(572.5) |
| **Maternal pre-pregnancy BMI (kg/m<sup>2</sup>)** | 22.7(3.4) | 22.7(3.4) |
| **Maternal age at delivery (years)** | 29(4.5) | 29(4.5) |

\* Measured at 32-weeks' gestation.

### Sex-specific association of SEP with lipoprotein and lipid traits

At age 7y, lower maternal education was associated with higher small and medium HDL particle concentrations and lower large and very large HDL concentrations in both sexes with some confidence intervals spanning the null (Figure 2; full results in eTables 5-6). For instance, less than O-level maternal education was associated with 0.27 SD (95% confidence interval (CI): 0.11, 0.43) and 0.26 SD (95% CI: 0.10, 0.42) higher small HDL concentrations compared with degree level in females and males respectively. Lower maternal education was also associated with smaller HDL and low-density lipoprotein (LDL) particle size and effect sizes were similar in females and males. Among females, lower maternal education was associated with higher concentrations of all very-low-density lipoproteins (VLDL), higher remnant and VLDL cholesterol concentrations and higher HDL, LDL, total and VLDL triglycerides and apolipoprotein B concentrations, albeit with some confidence intervals spanning the null. In contrast, among males, there was little evidence of associations with these lipoprotein and lipid traits at 7y. For instance, less than O-level maternal education was associated with 0.16 SD (95% CI: 0.01, 0.30) higher very small VLDL concentrations among females and -0.02 SD (95% CI: -0.16, 0.13) among males at 7y.

**Figure 2:**
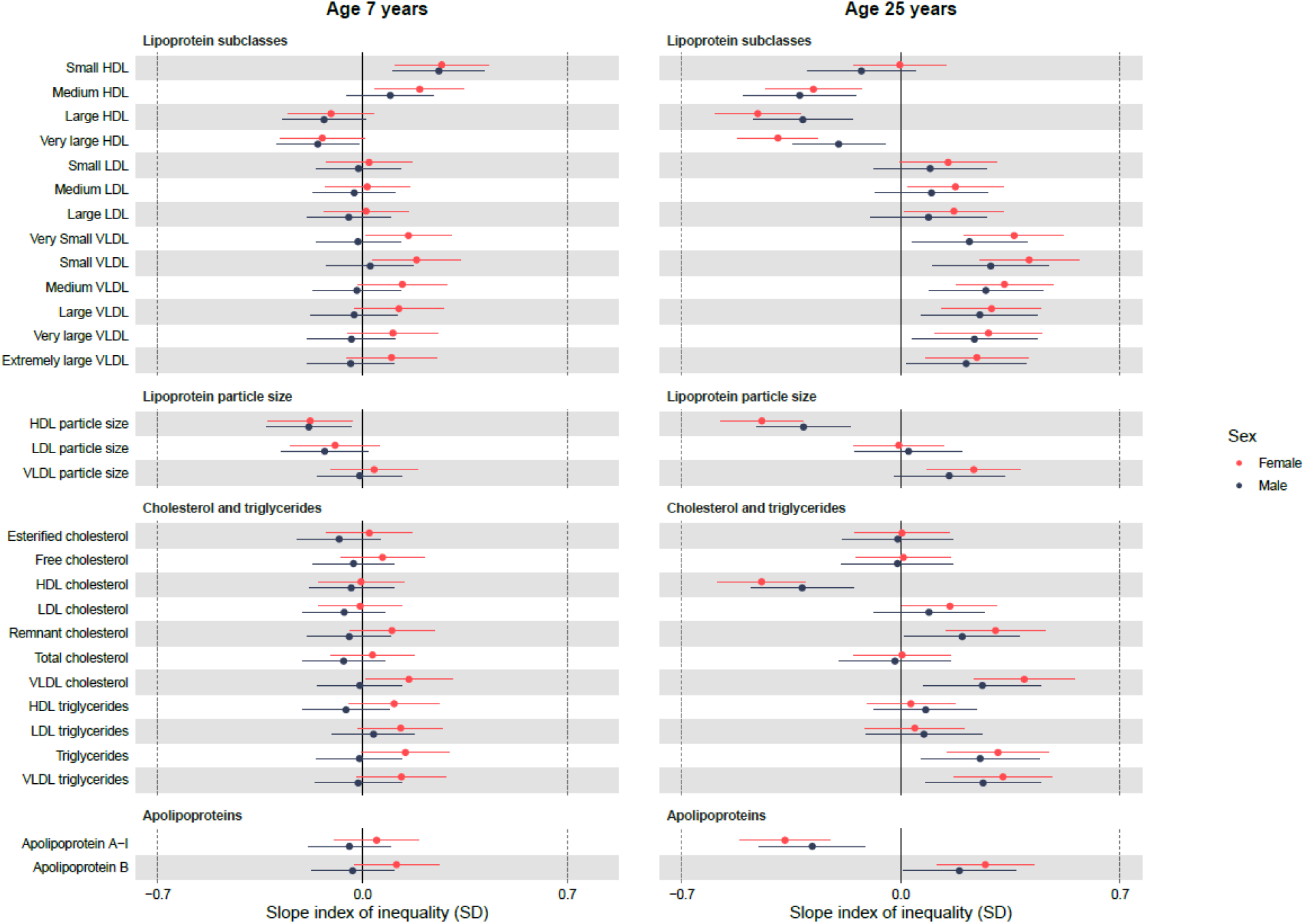
Sex-specific associations of maternal education with lipoprotein and lipid concentrations at 7y and 25y, estimated from multilevel models. Slope index of inequality represents the mean difference in SDs of the outcome between the individuals of lowest and highest socioeconomic position on the hypothetical underlying continuous distribution of maternal education. Results shown are standardised mean differences in metabolic traits comparing lowest maternal education (less than O-level) with highest maternal education (degree level) with whiskers representing 95% confidence intervals. HDL, high-density lipoprotein; LDL, low-density lipoprotein; SD, standard deviation; VLDL, very-low-density lipoprotein.

By 25y, associations of lower maternal education with higher VLDL particle concentrations, remnant and VLDL cholesterol concentrations, total and VLDL triglyceride concentrations and apolipoprotein B concentrations persisted and somewhat strengthened among females while associations of lower maternal education with higher concentrations of these traits emerged among males. For instance, less than O-level maternal education was associated with 0.36 SD (95% CI: 0.20, 0.52) and 0.22 SD (95% CI: 0.04, 0.40) higher very small VLDL concentrations at 25y among females and males respectively. By 25y, lower maternal education was associated with higher LDL particle and LDL cholesterol concentrations, larger VLDL particle size and lower HDL cholesterol and apolipoprotein A-1 concentrations, although many confidence intervals spanned the null among males. Associations of lower maternal education with lower medium HDL concentrations emerged while associations with lower large and very large HDL concentrations and smaller HDL particle size strengthened in both females and males. At 25y, there was little evidence of associations of maternal education with LDL particle size and free, esterified, and total cholesterol concentrations in both sexes.

Mean change in concentrations by maternal education, estimated from multilevel models, is shown in Table 2 (full results in eTable 7). Most trait concentrations decreased from 7y to 25y among females with degree level maternal education except for HDL and LDL particle concentrations and HDL cholesterol, HDL triglycerides and apolipoprotein A-1 which increased from 7y to 25y. Lower maternal education was associated with smaller decreases in very small, small and medium VLDL concentrations, apolipoprotein B concentrations and LDL, remnant, and VLDL cholesterol, smaller increases in HDL particle concentrations and larger increases in LDL particle concentrations among females. Lower maternal education was associated with decreasing HDL particle size and decreasing apolipoprotein A-1 and HDL cholesterol concentrations. Among males with degree level maternal education, all trait concentrations decreased from 7y to 25y except for small and medium HDL, all LDL particle concentrations, medium VLDL particle concentrations, and VLDL particle size which increased over time. Lower maternal education was associated with smaller increases in small and medium HDL concentrations and smaller decreases in VLDL concentrations, remnant and VLDL cholesterol, triglycerides and apolipoprotein B concentrations. Lower maternal education was associated with larger decreases in HDL particle size, HDL cholesterol and apolipoprotein A-1 concentrations.

**Table 2:** Mean change in lipoprotein and lipid concentrations between 7y and 25y in SD units, by maternal education, estimated from multilevel models.

|  | Females |  | Males |  |
| --- | --- | --- | --- | --- |
|  | Mean change (95% CI) in concentration in degree level maternal education | Mean difference (95% CI) in change in less than O-level maternal education | Mean change (95% CI) in concentration in degree level maternal education | Mean difference (95% CI) in change in less than O-level maternal education |
| **Lipoprotein subclasses** |  |  |  |  |
| Small HDL | 3.54 (3.33,3.75) | -0.28 (-0.64,0.09) | 3.07 (2.83,3.30) | -0.56 (-0.98,-0.14) |
| Medium HDL | 4.39 (4.16,4.61) | -0.90 (-1.29,-0.50) | 2.65 (2.39,2.90) | -0.90 (-1.37,-0.44) |
| Large HDL | 1.63 (1.46,1.80) | -0.90 (-1.21,-0.60) | -0.67 (-0.87,-0.48) | -0.56 (-0.91,-0.21) |
| Very large HDL | -0.15 (-0.28,-0.02) | -0.53 (-0.76,-0.30) | -1.87 (-2.01,-1.72) | -0.19 (-0.44,0.07) |
| Small LDL | 0.38 (0.23,0.53) | 0.23 (-0.04,0.50) | 0.55 (0.38,0.72) | 0.17 (-0.14,0.48) |
| Medium LDL | 0.80 (0.65,0.94) | 0.26 (0.01,0.51) | 0.98 (0.82,1.14) | 0.18 (-0.11,0.47) |
| Large LDL | 0.80 (0.65,0.94) | 0.25 (-0.004,0.49) | 0.82 (0.66,0.98) | 0.18 (-0.10,0.47) |
| Very Small VLDL | -0.94 (-1.08,-0.79) | 0.40 (0.15,0.65) | -0.69 (-0.84,-0.53) | 0.35 (0.07,0.64) |
| Small VLDL | -1.07 (-1.20,-0.94) | 0.33 (0.11,0.56) | -0.28 (-0.43,-0.14) | 0.34 (0.08,0.59) |
| Medium VLDL | -0.51 (-0.63,-0.38) | 0.24 (0.02,0.45) | 0.19 (0.06,0.32) | 0.33 (0.09,0.56) |
| Large VLDL | -0.55 (-0.66,-0.44) | 0.14 (-0.05,0.34) | -0.10 (-0.22,0.02) | 0.26 (0.05,0.47) |
| Very large VLDL | -0.72 (-0.82,-0.62) | 0.08 (-0.10,0.25) | -0.40 (-0.51,-0.29) | 0.19 (0.003,0.37) |
| Extremely large VLDL | -0.75 (-0.85,-0.65) | 0.05 (-0.12,0.22) | -0.44 (-0.54,-0.34) | 0.17 (-0.01,0.35) |
| **Lipoprotein particle size** |  |  |  |  |
| HDL particle size | -0.02 (-0.14,0.11) | -0.57 (-0.79,-0.34) | -1.69 (-1.83,-1.55) | -0.34 (-0.59,-0.08) |
| LDL particle size | -0.12 (-0.26,0.03) | 0.08 (-0.17,0.34) | -0.69 (-0.85,-0.53) | 0.17 (-0.12,0.45) |
| VLDL particle size | -0.49 (-0.60,-0.38) | 0.17 (-0.02,0.35) | 0.16 (0.05,0.27) | 0.15 (-0.05,0.34) |
| **Cholesterol and triglycerides** |  |  |  |  |
| Esterified cholesterol | -0.66 (-0.78,-0.54) | -0.02 (-0.24,0.20) | -0.94 (-1.08,-0.81) | 0.07 (-0.18,0.31) |
| Free cholesterol | -0.56 (-0.68,-0.44) | -0.06 (-0.27,0.16) | -0.87 (-1.00,-0.73) | 0.02 (-0.22,0.26) |
| HDL cholesterol | 0.40 (0.26,0.53) | -0.72 (-0.96,-0.48) | -1.23 (-1.38,-1.07) | -0.48 (-0.75,-0.20) |
| LDL cholesterol | -0.31 (-0.43,-0.19) | 0.21 (-0.0001,0.42) | -0.07 (-0.20,0.06) | 0.18 (-0.06,0.42) |
| Remnant cholesterol | -1.58 (-1.70,-1.46) | 0.28 (0.07,0.50) | -1.14 (-1.28,-1.01) | 0.29 (0.05,0.54) |
| Total cholesterol | -0.64 (-0.77,-0.52) | -0.03 (-0.25,0.19) | -0.91 (-1.04,-0.77) | 0.04 (-0.21,0.29) |
| VLDL cholesterol | -1.76 (-1.87,-1.64) | 0.30 (0.09,0.51) | -1.13 (-1.26,-1.00) | 0.31 (0.08,0.54) |
| HDL triglycerides | 0.48 (0.35,0.61) | -0.06 (-0.30,0.17) | -0.23 (-0.37,-0.09) | 0.17 (-0.09,0.42) |
| LDL triglycerides | -0.26 (-0.37,-0.15) | -0.09 (-0.28,0.09) | -0.74 (-0.86,-0.63) | 0.02 (-0.18,0.23) |
| Triglycerides | -0.68 (-0.79,-0.56) | 0.15 (-0.05,0.35) | -0.30 (-0.43,-0.18) | 0.25 (0.04,0.47) |
| VLDL triglycerides | -0.73 (-0.84,-0.61) | 0.19 (-0.01,0.38) | -0.15 (-0.27,-0.03) | 0.27 (0.05,0.49) |
| **Apolipoproteins** |  |  |  |  |
| Apolipoprotein A-I | 0.51 (0.36,0.67) | -0.71 (-0.98,-0.44) | -0.95 (-1.12,-0.78) | -0.46 (-0.77,-0.15) |
| Apolipoprotein B | -0.82 (-0.94,-0.69) | 0.23 (0.02,0.45) | -0.32 (-0.46,-0.19) | 0.27 (0.03,0.52) |

Results shown are standardised mean change and mean difference in metabolic traits. The intercept, spline periods and coefficients for the slope index of inequality from multilevel models were used to estimate the mean change in concentration for degree level maternal education and mean difference for less than O-level. CI, confidence interval; HDL, high-density lipoprotein; LDL, low-density lipoprotein; VLDL, very-low-density lipoprotein.

### Sex-specific association of SEP with amino acids, fatty acids and other metabolic traits

At 7y, lower maternal education was associated with lower concentrations of all amino acids in both sexes, except glutamine (Figure 3; full results in eTable 5-6). Lower maternal education was also associated with lower concentrations of most fatty acids among males, except for fatty acid chain length and monounsaturated fatty acids. Among females, lower maternal education was associated with lower concentrations of conjugated linoleic acid, omega-3, omega-6 and polyunsaturated fatty acids though confidence intervals spanned the null for omega-6 and polyunsaturated fatty acids and there was little evidence of associations with fatty acid chain length, linoleic acid, and saturated and total fatty acids. Except for acetate, lower maternal education was associated with higher concentrations of all other metabolites, though confidence intervals spanned the null for creatinine and glucose among males and there was little evidence of associations with GlycA among males. For instance, less than O-level maternal education was associated with 0.21 SD (95% CI: 0.06, 0.37) higher concentrations of GlycA among females and 0.03 SD (95% CI: -0.13, 0.18) among males.

**Figure 3:**
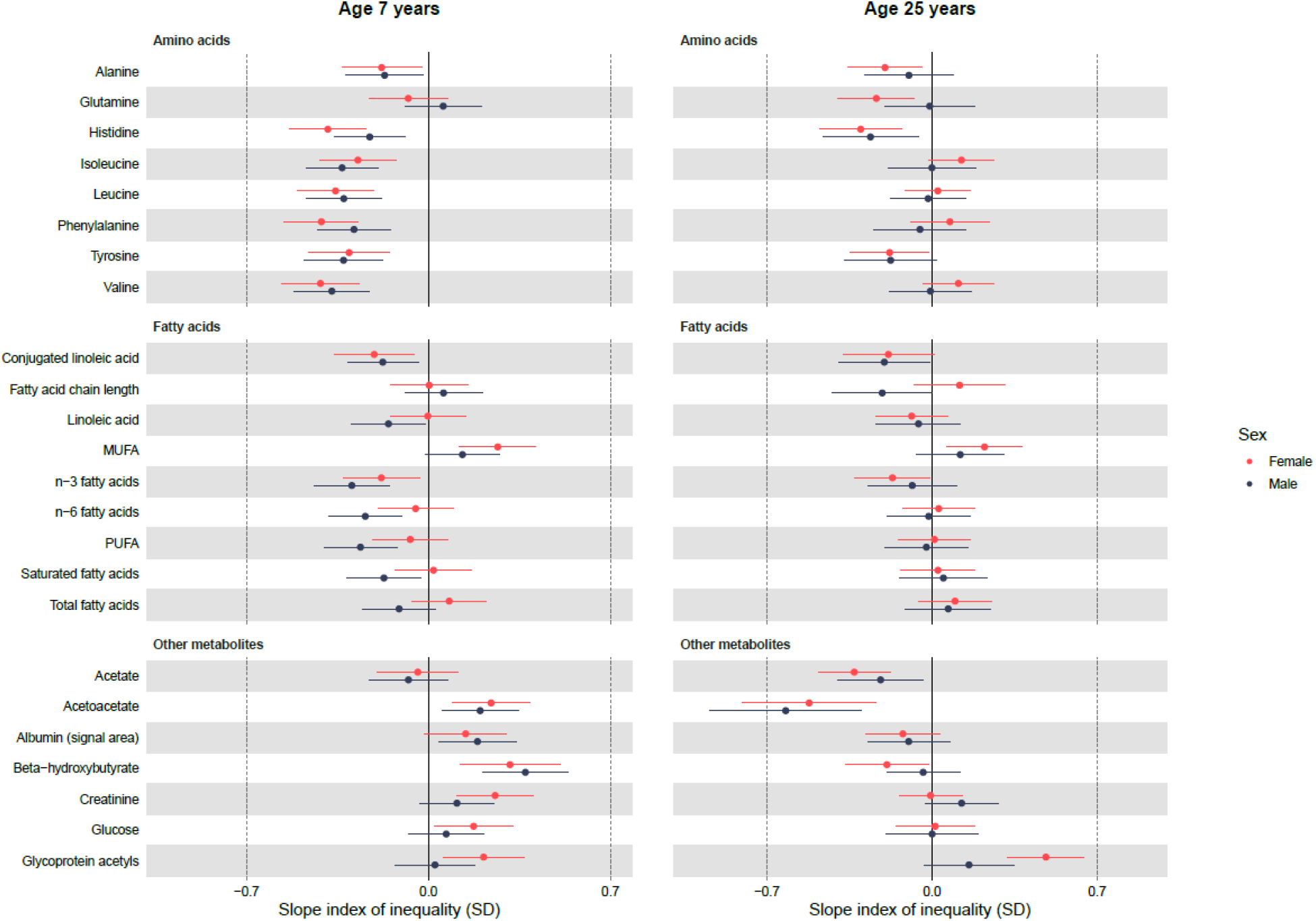
Sex-specific associations of maternal education with amino acids, fatty acids and other metabolic at 7y and 25y, estimated from multilevel models. Slope index of inequality represents the mean difference in SDs of the outcome between the individuals of lowest and highest socioeconomic position on the hypothetical underlying continuous distribution of maternal education. Results shown are standardised mean differences in metabolic traits comparing lowest maternal education (less than O-level) with highest maternal education (degree level) with whiskers representing 95% confidence intervals. MUFA, monounsaturated fatty acids; PUFA, polyunsaturated fatty acids; SD, standard deviation; VLDL, very-low-density lipoprotein. *Conjugated linoleic acid and fatty acid chain length are modelled up to 18y only.

By 25y, associations of lower maternal education with lower concentrations of several amino acids (isoleucine, leucine, phenylalanine and valine) had weakened in both sexes (Figure 3; full results in eTable 5-6). Associations with lower histidine, tyrosine and alanine concentrations persisted at 25y in both sexes as did associations with lower glutamine concentrations in females but not males. Similarly, associations of lower maternal education with lower concentrations of several fatty acids at 25y also weakened (omega-3, omega-6, polyunsaturated, saturated, and total fatty acids). Associations of lower maternal education with lower conjugated linoleic acid, linoleic acid and higher monounsaturated fatty acid at 7y remained broadly similar at 25y. By 25y, associations of lower maternal education with higher GlycA concentrations strengthened among females and emerged among males. For instance, less than O-level maternal education was associated with 0.48 SD (95% CI: 0.32, 0.65) and 0.16 SD (95% CI: -0.03, 0.35) higher GlycA concentrations in females and males respectively. Associations of maternal education with acetoacetate changed direction such that lower maternal education was associated with lower concentrations by 25y, as did associations with albumin and beta-hydroxybutate, though confidence intervals spanned the null.

Mean absolute change in concentrations by maternal education, estimated from multilevel models, is shown in Table 3 (full results in eTable 7). All trait concentrations decreased between age 7y and 25y among females with degree level maternal education except for alanine, phenylalanine, fatty acid chain length, albumin, beta-hydroxybutyrate and creatinine which increased over time. Lower maternal education was associated with smaller decreases in several amino acids, smaller increases in beta-hydroxybutate and creatinine, larger decreases in acetoacetate and with increases in GlycA concentrations. Most trait concentrations also decreased between 7y and 25y among males with degree level maternal education except for alanine, leucine, phenylalanine, valine, fatty acid chain length, albumin, beta-hydroxybutyrate and creatinine which increased over the period. Lower maternal education was associated with larger increases in these amino acid concentrations, smaller increases in albumin and beta-hydroxybutyrate concentrations and larger decreases in acetoacetate concentrations.

**Table 3:** Mean change in amino acids, fatty acids and other metabolic concentrations between 7y and 25y in females, by maternal education, estimated from multilevel models.

|  | Females |  | Males |  |
| --- | --- | --- | --- | --- |
|  | Mean change (95% CI) in concentration in degree level maternal education | Mean difference (95% CI) in change in less than O-level maternal education | Mean change (95% CI) in concentration in degree level maternal education | Mean difference (95% CI) in change in less than O-level maternal education |
| **Amino acids** |  |  |  |  |
| Alanine | 0.64 (0.52,0.76) | -0.001 (-0.21,0.21) | 0.84 (0.71,0.97) | 0.08 (-0.14,0.30) |
| Glutamine | -2.40 (-2.54,-2.27) | -0.21 (-0.45,0.03) | -0.96 (-1.11,-0.80) | -0.07 (-0.34,0.20) |
| Histidine | -2.17 (-2.28,-2.07) | 0.20 (0.02,0.38) | -1.69 (-1.79,-1.58) | 0.06 (-0.12,0.25) |
| Isoleucine | -0.66 (-0.75,-0.56) | 0.35 (0.18,0.51) | -0.05 (-0.15,0.05) | 0.33 (0.15,0.51) |
| Leucine | -0.37 (-0.48,-0.27) | 0.38 (0.20,0.56) | 0.42 (0.31,0.54) | 0.31 (0.12,0.50) |
| Phenylalanine | 0.28 (0.18,0.39) | 0.47 (0.29,0.65) | 0.60 (0.48,0.71) | 0.25 (0.06,0.44) |
| Tyrosine | -1.84 (-1.94,-1.73) | 0.20 (0.01,0.38) | -1.49 (-1.60,-1.38) | 0.22 (0.03,0.41) |
| Valine | -0.59 (-0.69,-0.48) | 0.50 (0.32,0.69) | 0.23 (0.11,0.34) | 0.37 (0.17,0.56) |
| **Fatty acids** |  |  |  |  |
| Conjugated linoleic acid | -0.55 (-0.67,-0.43) | 0.08 (-0.12,0.28) | -0.51 (-0.62,-0.40) | 0.03 (-0.15,0.22) |
| Fatty acid chain length | 0.42 (0.28,0.55) | 0.11 (-0.12,0.34) | 0.61 (0.47,0.74) | -0.26 (-0.50,-0.02) |
| Linoleic acid | -1.17 (-1.29,-1.05) | -0.10 (-0.31,0.11) | -1.37 (-1.50,-1.23) | 0.08 (-0.15,0.32) |
| MUFA | -0.47 (-0.60,-0.34) | -0.01 (-0.23,0.22) | -0.49 (-0.63,-0.35) | 0.01 (-0.24,0.26) |
| n-3 fatty acids | -0.62 (-0.75,-0.50) | -0.01 (-0.22,0.21) | -0.69 (-0.82,-0.56) | 0.20 (-0.04,0.44) |
| n-6 fatty acids | -1.20 (-1.33,-1.08) | 0.09 (-0.13,0.30) | -1.51 (-1.64,-1.38) | 0.23 (-0.01,0.47) |
| PUFA | -1.19 (-1.31,-1.07) | 0.08 (-0.13,0.30) | -1.47 (-1.61,-1.34) | 0.23 (-0.01,0.48) |
| Saturated fatty acids | -1.36 (-1.48,-1.24) | 0.01 (-0.20,0.21) | -1.62 (-1.75,-1.49) | 0.22 (-0.01,0.45) |
| Total fatty acids | -1.12 (-1.25,-1.00) | 0.04 (-0.18,0.26) | -1.34 (-1.48,-1.21) | 0.20 (-0.05,0.44) |
| **Other metabolites** |  |  |  |  |
| Acetate | -0.50 (-0.60,-0.40) | -0.13 (-0.31,0.04) | -0.41 (-0.51,-0.31) | -0.04 (-0.22,0.14) |
| Acetoacetate | -0.45 (-0.56,-0.34) | -0.39 (-0.58,-0.21) | -0.39 (-0.50,-0.28) | -0.38 (-0.57,-0.19) |
| Albumin | 0.24 (0.08,0.41) | -0.34 (-0.63,-0.05) | 1.40 (1.23,1.58) | -0.35 (-0.66,-0.04) |
| Beta-hydroxybutyrate | 1.38 (1.21,1.55) | -0.54 (-0.83,-0.25) | 1.20 (1.07,1.34) | -0.42 (-0.66,-0.17) |
| Creatinine | 3.07 (2.94,3.21) | -0.27 (-0.50,-0.03) | 5.02 (4.88,5.17) | 0.09 (-0.17,0.35) |
| Glucose | -0.49 (-0.59,-0.39) | -0.17 (-0.34,0.01) | -0.33 (-0.44,-0.23) | -0.07 (-0.25,0.11) |
| Glycoprotein acetyls | -0.28 (-0.42,-0.14) | 0.40 (0.16,0.65) | -0.13 (-0.29,0.02) | 0.17 (-0.10,0.45) |

Results shown are standardised mean change and mean difference in metabolic traits. The intercept, spline periods and coefficients for the slope index of inequality from multilevel models were used to estimate the mean change in concentration for degree level maternal education and mean difference for less than O-level. CI, confidence interval; MUFA, monounsaturated fatty acids; PUFA, polyunsaturated fatty acids; VLDL, very-low-density lipoprotein. \*Conjugated linoleic acid and fatty acid chain length are modelled up to 18y only.

### Additional and sensitivity analyses

Participants included in analyses of parental education and household social class are detailed in eFigures 1-2. Overall, associations of paternal education and household social class with metabolic traits followed largely similar patterns to those observed for maternal education (eFigures 3-6, eTables 8-11). However, at age 25y, associations of paternal education among males were weaker than those observed for maternal education. Results from multi-level models with maternal education included as a categorial variable (as opposed to calculating the SII as a summary measure) are displayed in eFigures 7-10. Among females, similar patterns of associations with several lipid and lipoprotein traits were observed across categories of lower maternal education at age 7y. In contrast, there was little evidence of associations in males. By age 25y, such associations strengthened among females and there was evidence of a socioeconomic gradient in associations across lower maternal education categories. Among males at 25y, associations emerged particularly for the two lowest maternal education categories (less than O-level and O-level education).

Results from the inverse probability weighted sensitivity analysis were not appreciably different from main findings (eFigures 11-12).

## Discussion

In this large prospective cohort study including three indicators of early life SEP and repeated measurements of 148 molecular measures of systemic metabolism from 7y to 25y, we found socioeconomic inequalities in causal atherogenic lipids and other key metabolic traits such as markers of inflammation whereby disadvantaged SEP was associated with a more adverse metabolic profile. Socioeconomic inequalities in these causal atherogenic lipids and other key metabolic traits begin in childhood and strengthen in adolescence among females but only emerge in adolescence among males, leading to wider socioeconomic inequalities among females compared with males by 25y. Our findings suggest that childhood and adolescence are important periods for the embodiment of socioeconomic disadvantage via systemic metabolism in both sexes and are also important periods for the emergence of sex differences in socioeconomic inequalities in cardiometabolic risk to the detriment of females. Prevention of socioeconomic inequalities in cardiometabolic risk requires a life course approach that begins at the earliest opportunity in the life course especially among females.

Socioeconomic disadvantage leads to an increased risk of CVD (4,5) and more recently has been linked with a metabolic profile predictive of CVD that appears to emerge in early life (9). This recent sex-combined analysis of ALSPAC data examined associations of father’s occupation with a subset of the metabolic traits included in our analysis and demonstrated that children of manual workers had a more adverse metabolic profile at age 7y and age 17y compared to children of non-manual workers. Our results build on these findings and suggest that much of the observed pattern in the previous analysis may be driven by associations of SEP with metabolic traits in females given overall evidence of weaker associations in males. Furthermore, we applied a formal trajectory modelling approach to extend results beyond adolescence into early adulthood and included a larger profile of metabolic traits, demonstrating that by age 25y socioeconomic inequalities across several indicators of early life SEP are evident in key atherogenic traits levels known to causally affect CVD risk (33) and biomarkers predictive for cardiometabolic diseases (34,35). For instance, apolipoprotein B has been identified as the predominant lipoprotein trait that plays a fundamental role in the aetiology of coronary heart disease (36). Among females, we found higher apolipoprotein B concentrations among those with disadvantaged SEP at 7y which strengthened by 25y, while higher concentrations only began to emerge among males with disadvantaged SEP during adolescence and early adulthood. Similarly, we found higher particle concentrations of GlycA at 7y, a biomarker for chronic inflammation that predicts risk of incident CVD (34,35), among those with disadvantaged SEP, particularly among females, and socioeconomic inequalities widened between age 7y and 25y. Our findings also demonstrated that while disadvantaged SEP was associated with levels of amino acids and some fatty acids among females and males at age 7y, most associations weakened over time with little evidence of associations by age 25y. For instance, disadvantaged SEP was associated with lower levels of branched-chain amino acids valine and leucine at age 7y but not 25y. Early life SEP influences diet in childhood (37) which may mediate the observed associations at age 7y but not at 25y.

While few studies have explored sex-specific SEP associations with detailed markers of CVD risk in early life, our findings are similar to a previous study also conducted in ALSPAC which showed wider socioeconomic inequalities (using maternal education) in adiposity, blood pressure and apolipoprotein B at age 10 years in females compared with males (10). Another ALSPAC study examining socioeconomic inequalities in trajectories of adiposity from birth to 10 years among females and males found associations between maternal education and adiposity emerging by 4 years of age in both sexes and strengthening with increasing age, particularly in females (7). Overall, socioeconomic inequality in adiposity was wider among females than males. Taken together, these findings suggest that adiposity may mediate associations of SEP and metabolic traits given the similarity with the patterns of associations observed in our study. Recent analyses, also conducted in ALSPAC, have shown that higher adiposity at age 10 years is associated with metabolic traits at age 18 years, particularly higher VLDL and LDL cholesterol, lower HDL cholesterol, higher triglycerides, and higher concentrations of GlycA (38). Furthermore, adiposity has been shown to be more strongly associated with key metabolic traits in males between the ages of 15 and 25 years compared with females (39) and may therefore explain why we observed associations of SEP with metabolic traits emerging later in males given that the effect of SEP on adiposity is weaker in males (10). However, reasons for sex differences in the patterning of SEP associations with cardiometabolic traits largely remain unexplained. Differential socioeconomic patterning of physical activity in females and males has been posited as a potential mediator in the sex-specific association of SEP and adiposity. While physical activity is associated with metabolic traits in adolescence, associations are small in magnitude and likely short lived (40) and therefore it is unlikely to be a strong mediator in the sex-specific associations observed here between SEP and metabolic traits at 25 years.

Sex differences in the relationship between SEP and CVD events are well established with stronger associations observed for females compared with males but the mechanisms underpinning these differences are not well understood (12). Our findings suggest that the excess cardiovascular risk conferred by disadvantaged SEP in females may begin early in life and strengthen through adolescence into early adulthood. Overall, SEP was more consistently associated with CVD-relevant metabolites among females in our study such as apolipoprotein B (36) and GlycA (34,35), in contrast to weaker associations among males in early adulthood particularly for paternal education. Thus, our findings suggest that sex differences in socioeconomic inequalities in CVD may not be solely explained by differential identification of CVD risk and access to secondary and tertiary prevention in mid-life among women (12). However, further research is required to determine whether these associations track into mid-life or strengthen over time and whether such circulating cardiometabolic traits mediate the pathway from childhood SEP to CVD events. Our findings highlight the importance of sex-specific analyses in future work investigating mediators of associations between childhood SEP and CVD risk.

### Strengths and Limitations

There are several strengths to our study including the use of 148 molecular metabolic trait concentrations from a targeted metabolomics platform measured at four occasions from childhood to early adulthood to characterise early life course trajectories in a relatively large sample and the use of three different indicators of SEP across the early life course. We used multilevel models which take account of clustering of repeated measures within individuals and the correlation between measures over time. Multilevel models also allow for the inclusion of all participants with at least one measure of a metabolic trait throughout the follow-up period, thereby minimising potential for selection bias and increasing our sample size. However, there are also limitations: the possibility of selection bias remains given that participants included in our analysis were more likely to have an advantaged SEP compared with those excluded due to missing data. We conducted a sensitivity analysis using inverse probability weighting to account for the higher probability of inclusion of participants with higher SEP and results were similar to our main findings. Multilevel models include all participants with at least one measure of a metabolic trait under the missing at random (MAR) assumption, thus we assume that within categories of SEP there are no systematic differences between observed and missing values. There was differential loss-to-follow-up by SEP with higher attrition in those with disadvantaged SEP, particularly among males. Should the MAR assumption be violated, and missing values are more adverse than the observed values of metabolic traits, our findings may be biased. ALSPAC is a contemporary prospective cohort study, however SEP data refers to childhood SEP in the early 1990s. The meaning of SEP indicators such as education and household social class change over time and as such likely cohort effects need to be considered in the interpretation of findings (41).

## Conclusion

Socioeconomic inequalities in causal atherogenic lipids and other key metabolic traits such as markers of inflammation begin in childhood and strengthen in adolescence among females but only emerge in adolescence among males, leading to wider socioeconomic inequalities among females compared with males by 25y. Our findings suggest that childhood and adolescence are important periods for the embodiment of socioeconomic disadvantage in both sexes and are also important periods for the emergence of sex differences in socioeconomic inequalities in cardiometabolic risk to the detriment of females. Prevention of socioeconomic inequalities in cardiovascular disease risk requires a life course approach that begins at the earliest opportunity in the life course especially among females.

## Data Availability

Individual-level ALSPAC data are available following an application. This process of managed access is detailed at www.bristol.ac.uk/alspac/researchers/access. Cohort details and data descriptions for ALSPAC are publicly available at the same web address.

## Acknowledgements

We are extremely grateful to all the families who took part in this study, the midwives for their help in recruiting them, and the whole ALSPAC team, which includes interviewers, computer and laboratory technicians, clerical workers, research scientists, volunteers, managers, receptionists and nurses.

