## Supplementary Materials for "Sex-specific associations of childhood socioeconomic position and trajectories of metabolic traits across early life: prospective cohort study"

**Supplementary Material**

**eMethods 1:** Details of quality control

**eMethods 2:** Model selection

**eTable 1:** Metabolic trait subclass, name, units and multilevel model details for each

**eMethods 3:** Characteristics of included vs. excluded participants

**eTable 2:** Results from likelihood ratio test examining linearity of association between maternal education and metabolic traits at each age by sex

**eTable 3:** Characteristics at birth of the mothers of children included in models compared with those excluded due to missing exposure or outcome data

**eTable 4:** Number of participants with measures at each time point

**eTable 5:** Associations of maternal education with concentrations of 148 metabolic traits at age 7 years and 25 years in SD units, estimated from multilevel models

**eTable 6:** Associations of maternal education with concentrations of 148 metabolic traits at age 7 years and 25 years in original units, estimated from multilevel models

**eTable 7:** Mean rate of change in concentrations of 148 metabolic traits by maternal education in SD units, estimated from multilevel models

**eFigure 1:** Flowchart of participants included in paternal education analyses

**eFigure 2:** Flowchart of participants included in household social class analyses

**eFigure 3:** Associations of paternal education with lipoprotein and lipid concentrations at 7y and 25y, estimated from multilevel models

**eTable 8:** Mean change in lipoprotein and lipid concentrations between 7y and 25y, by paternal education, estimated from multilevel models

**eFigure 4:** Associations of paternal education with amino acids, fatty acids and other metabolic concentrations at 7y and 25y, estimated from multilevel models

**eTable 9:** Mean change in amino acids, fatty acids and other metabolic concentrations between 7y and 25y in females, by paternal education, estimated from multilevel models

**eFigure 5:** Associations of household social class with lipoprotein and lipid concentrations at 7y and 25y, estimated from multilevel models

**eTable 10:** Mean change in lipoprotein and lipid concentrations between 7y and 25y, household social class, estimated from multilevel models

**eFigure 6:** Associations of household social class with amino acids, fatty acids and other metabolic concentrations at 7y and 25y, estimated from multilevel models

**eTable 11:** Mean change in amino acids, fatty acids and other metabolic concentrations between 7y and 25y in females, by household social class, estimated from multilevel models

**eFigure 7:** Associations of lower maternal education categories (compared with degree level) with lipoprotein and lipid concentrations at 7y and 25y in females, estimated from multilevel models

**eFigure 8:** Associations of lower maternal education categories (compared with degree level) with amino acids, fatty acids and other metabolic concentrations at 7y and 25y in females, estimated from multilevel models

**eFigure 9:** Associations of lower maternal education categories (compared with degree level) with lipoprotein and lipid concentrations at 7y and 25y in males, estimated from multilevel models

**eFigure 10:** Associations of lower maternal education categories (compared with degree level) with amino acids, fatty acids and other metabolic concentrations at 7y and 25y in males, estimated from multilevel models

**eFigure 11:** Weighted associations of maternal education with lipoprotein and lipid concentrations at 7y and 25y, estimated from multilevel models

**eFigure 12:** Weighted associations of maternal education with amino acids, fatty acids and other metabolic concentrations at 7y and 25y, estimated from multilevel models

**eMethods 1: Details of quality control**

*Laboratory*

For each sample, the NMR spectra were analysed for absolute metabolite quantification (molar concentration) in automated fashion. A ridge regression model was applied for quantification of each metabolite in order to overcome the problems of heavily overlapping spectral data. Quantification of lipoprotein lipid data was performed by calibrating against high performance liquid chromatography methods, and then individually cross-validated against NMR-independent lipid data. Low-molecular-weight metabolites, as well as lipid extract measures, were quantified as mmol/l based on regression modelling calibrated against a set of manually fitted metabolite measures. The calibration data was quantified based on iterative line-shape fitting analysis using PERCH NMR software (PERCH Solutions Ltd., Kuopio, Finland). Absolute quantification could not be directly established for the lipid extract measures due to experimental variation in the lipid extraction protocol. Therefore, serum extract metabolites have been scaled via the total cholesterol as quantified from the native serum LIPO spectrum. We have previously shown strong correlation between the NMR and clinical chemistry measures that are available from both methods.

*Data preparation*

Prior to statistical analysis, preparation of metabolomics data was performed for each occasion separately using the R package metaboprep (<https://github.com/MRCIEU/metaboprep>) (version 0.0.1). QC was performed excluding the derived metabolomics measures from missingness and clustering. Briefly, individuals, and then metabolites, with high missingness (>=80%) were removed. Missingness was then re-calculated for individuals and metabolites, with removal based on >=20% missingness. Individuals were then removed based on total sum abundance, considering outliers as > 5 standard deviations away from the mean. Outliers are identified as being > 5 standard deviations away from the mean of principal component 1 and 2 and were excluded.

**eMethods 2:**  **Model selection**

Linear splines allow knot points to be fit at different ages to derive periods in which change is approximately linear. To select the optimal linear spline model for 144 trait concentrations measured from 7y to 25y, a series of models were ran including model 1: a model with two linear spline periods each (7y to 18y and 18y to 25y), model 2: a second model with two linear spline periods (7y to 15y and 15y to 25y), and model 3: a single slope model (a single age term which assumed constant change from 7y to 25y). Linear spline periods were chosen to reflect ages in whole years that were closest to mean age at clinics and hence where the density of measures was greatest; note that the same process was carried out to select models for the four traits with measures only available to 18y with a model with two linear spline periods (7y to 15y and 15y to 18y) and single slope (7y to 18y) being compared. For each trait and model, Akaike’s Information Criterion (AIC) was examined as an indicator of model fit with lower AIC values indicative of better model fit. Upon selection of the best fitting model based on AIC, observed and predicted values of models were examined to further assess model fit. Age (in years) was centred at the first available measure (7y). For each risk factor, all participants with at least one measure of the risk factor were included in each multilevel model, under a missing at random (MAR) assumption, to minimise selection bias.

Following the above process of model comparisons, the final models selected for 68 of 144 outcomes (see eTable 1) had two linear spline periods from 7-15 and 15-25, 75 of 144 outcomes had two linear spline periods from 7-18 and 18-25 and one outcome was a single slope model from 7y to 25y. For the four outcomes measured only up to 18y, final models selected had two linear spline periods from 7-15 and 15-18. Models were estimated with robust standard errors for both fixed effects and individual level random effects to account for skewed distributions in some traits. Unstructured variance-covariance matrices for the individual level random effects were used to estimate most trajectories; a complete list of traits, the optimal linear spline model selected and other model details including details of the variance-covariance matrix for each trait are shown in eTable 1. For 36 of 148 outcomes modelled (eTable 1), the covariance of the individual level random effects (level 2) were set to zero for some parameters to improve model convergence.

To explore the sex-specific associations between maternal education and metabolic traits, the slope index of inequality was estimated (30). A rank score of maternal education from 0 to 1 was created whereby the score for those in each category is the mid-point of the cumulative proportion of the participants in that category or lower. A three-way interaction term between sex, rank score of maternal education and the intercept and each linear spline period was included in all models. This allowed the sex-specific associations of maternal education with metabolic trajectories to be estimated from a single sex-combined model by using a linear combination of main effects and interaction effects for sex, rank score of maternal education, intercepts, and slopes. The coefficient for the maternal education rank score represents the slope index of inequality and can be interpreted as the mean difference in outcome between those in the lowest SEP level (less than O-level maternal education) compared to the highest SEP (degree level maternal education) on the hypothetical underlying continuous distribution of maternal education. Thus the model for each outcome took the form of metabolite_ij_ = (β_0_ + u_0j_) + β_1_*sex + (β_2_+ u_1j_)s_ij1_ + (β_3_+ u_2j_)s_ij2_ + (β_4_)s_ij1_*sex + (β_5_)s_ij2_*sex +  β_6_ *SEP + (β_7_)s_ij1_*SEP + (β_8_)s_ij2_*SEP +  β_9_*SEP*sex + (β_10_)s_ij1_*SEP*sex + (β_11_)s_ij2_*SEP*sex + e_ij_ where for person j at measurement occasion i; β_0_ represents the fixed effect coefficient for the average intercept in males with maternal degree level education, β_1_ represents the difference between the intercept for females compared with males, β_2_ and β_3_ represent fixed effect coefficients for the average linear slopes of each linear spline in males with maternal degree level education, s_ij_ represents the specific spline period, β_4_ and β_5_ represent the difference in the fixed effect coefficients for the average linear slopes of each linear spline in females with maternal degree level education compared with males, β_6_ represents the slope index of inequality in males, β_7_ and β_8_ represent the slope index of inequality for the average linear slopes of each linear spline in males, β_9_ represents the difference between the intercept for the slope index of inequality in females compared with males, β_10_ and β_11_ represent the difference in the slope index of inequality for the average linear slopes of each linear spline in females compared with males, u_0j_ to u_2j_ indicate person-specific (or individual level/level 2) random effects for the intercept and slopes respectively, and e_ij_ represents the occasion-specific residuals or measurement error which was allowed to vary by the intercept.

| **eTable 1: Metabolic trait subclass, name, units and multilevel model details for each** | | | | | |
| --- | --- | --- | --- | --- | --- |
| **Molecular class** | **Lipid, lipoprotein or metabolite name** | **Units** | **Knot** | **Level 2 variance** | **Level 1 variance** |
| **Extremely large VLDL** | Concentration of chylomicrons and extremely large VLDL particles | mol/l | 15 | Intercept & splines; unstructured | Intercept |
|  | Total lipids in chylomicrons and extremely large VLDL | mmol/l | 15 | Intercept & splines; matrix a = (1, 1, 1, 1, 1, 0) | Intercept |
|  | Phospholipids in chylomicrons and extremely large VLDL | mmol/l | 15 | Intercept & splines; unstructured | Intercept |
|  | Total cholesterol in chylomicrons and extremely large VLDL | mmol/l | 18 | Intercept & splines; matrix a = (1, 1, 1, 1, 1, 0) | Intercept |
|  | Cholesterol esters in chylomicrons and extremely large VLDL | mmol/l | 18 | Intercept & splines; matrix a = (1, 1, 1, 1, 1, 0) | Intercept |
|  | Free cholesterol in chylomicrons and extremely large VLDL | mmol/l | 15 | Intercept & splines; matrix a = (1, 1, 1, 1, 1, 0) | Intercept |
|  | Triglycerides in chylomicrons and extremely large VLDL | mmol/l | 15 | Intercept & splines; matrix a = (1, 1, 1, 1, 1, 0) | Intercept |
| **Very large VLDL** | Concentration of very large VLDL particles | mol/l | 15 | Intercept & splines; unstructured | Intercept |
|  | Total lipids in very large VLDL | mmol/l | 15 | Intercept & splines; unstructured | Intercept |
|  | Phospholipids in very large VLDL | mmol/l | 15 | Intercept & splines; unstructured | Intercept |
|  | Total cholesterol in very large VLDL | mmol/l | 15 | Intercept & splines; matrix a = (1, 1, 1, 1, 1, 0) | Intercept |
|  | Cholesterol esters in very large VLDL | mmol/l | 18 | Intercept & splines; unstructured | Intercept |
|  | Free cholesterol in very large VLDL | mmol/l | 15 | Intercept & splines; matrix a = (1, 1, 1, 1, 1, 0) | Intercept |
|  | Triglycerides in very large VLDL | mmol/l | 15 | Intercept & splines; unstructured | Intercept |
| **Large VLDL** | Concentration of large VLDL particles | mol/l | 15 | Intercept & splines; unstructured | Intercept |
|  | Total lipids in large VLDL | mmol/l | 15 | Intercept & splines; unstructured | Intercept |
|  | Phospholipids in large VLDL | mmol/l | 15 | Intercept & splines; unstructured | Intercept |
|  | Total cholesterol in large VLDL | mmol/l | 15 | Intercept & splines; unstructured | Intercept |
|  | Cholesterol esters in large VLDL | mmol/l | 15 | Intercept & splines; unstructured | Intercept |
|  | Free cholesterol in large VLDL | mmol/l | 15 | Intercept & splines; unstructured | Intercept |
|  | Triglycerides in large VLDL | mmol/l | 15 | Intercept & splines; unstructured | Intercept |
| **Medium VLDL** | Concentration of large VLDL particles | mol/l | 15 | Intercept & splines; unstructured | Intercept |
|  | Total lipids in small VLDL | mmol/l | 15 | Intercept & splines; unstructured | Intercept |
|  | Phospholipids in small VLDL | mmol/l | 15 | Intercept & splines; unstructured | Intercept |
|  | Total cholesterol in small VLDL | mmol/l | 15 | Intercept & splines; unstructured | Intercept |
|  | Cholesterol esters in small VLDL | mmol/l | 15 | Intercept & splines; unstructured | Intercept |
|  | Free cholesterol in small VLDL | mmol/l | 15 | Intercept & splines; unstructured | Intercept |
|  | Triglycerides in small VLDL | mmol/l | 15 | Intercept & splines; unstructured | Intercept |
| **Small VLDL** | Concentration of small VLDL particles | mol/l | 15 | Intercept & splines; unstructured | Intercept |
|  | Total lipids in small VLDL | mmol/l | 18 | Intercept & splines; unstructured | Intercept |
|  | Phospholipids in small VLDL | mmol/l | 15 | Intercept & splines; matrix a = (1, 1, 0, 1, 1, 1) | Intercept |
|  | Total cholesterol in small VLDL | mmol/l | 18 | Intercept & splines; unstructured | Intercept |
|  | Cholesterol esters in small VLDL | mmol/l | 18 | Intercept & splines; unstructured | Intercept |
|  | Free cholesterol in small VLDL | mmol/l | 18 | Intercept & splines; unstructured | Intercept |
|  | Triglycerides in small VLDL | mmol/l | 15 | Intercept & splines; unstructured | Intercept |
| **Very small VLDL** | Concentration of very small VLDL particles | mmol/l | 15 | Intercept & splines; matrix a = (1, 1, 0, 1, 1, 1) | Intercept |
|  | Total lipids in very small VLDL | mmol/l | 18 | Intercept & splines; unstructured | Intercept |
|  | Phospholipids in very small VLDL | mmol/l | 15 | Intercept & splines; matrix a = (1, 1, 0, 1, 1, 1) | Intercept |
|  | Total cholesterol in very small VLDL | mmol/l | 18 | Intercept & splines; unstructured | Intercept |
|  | Cholesterol esters in very small VLDL | mmol/l | 18 | Intercept & splines; unstructured | Intercept |
|  | Free cholesterol in very small VLDL | mmol/l | 18 | Intercept & splines; unstructured | Intercept |
|  | Triglycerides in very small VLDL | mmol/l | 18 | Intercept & splines; unstructured | Intercept |
| **IDI** | Concentration of IDL particles | mol/l | 18 | Intercept & splines; unstructured | Intercept |
|  | Total lipids in IDL | mmol/l | 18 | Intercept & splines; unstructured | Intercept |
|  | Phospholipids in IDL | mmol/l | 18 | Intercept & splines; unstructured | Intercept |
|  | Total cholesterol in IDL | mmol/l | 18 | Intercept & splines; unstructured | Intercept |
|  | Cholesterol esters in IDL | mmol/l | 18 | Intercept & splines; unstructured | Intercept |
|  | Free cholesterol in IDL | mmol/l | 18 | Intercept & splines; unstructured | Intercept |
|  | Triglycerides in IDL | mmol/l | 18 | Intercept & splines; unstructured | Intercept |
| **Large LDL** | Concentration of large LDL particles | mol/l | 18 | Intercept & splines; unstructured | Intercept |
|  | Total lipids in large LDL | mmol/l | 18 | Intercept & splines; unstructured | Intercept |
|  | Phospholipids in large LDL | mmol/l | 18 | Intercept & splines; unstructured | Intercept |
|  | Total cholesterol in large LDL | mmol/l | 18 | Intercept & splines; unstructured | Intercept |
|  | Cholesterol esters in large LDL | mmol/l | 18 | Intercept & splines; unstructured | Intercept |
|  | Free cholesterol in large LDL | mmol/l | 18 | Intercept & splines; unstructured | Intercept |
|  | Triglycerides in large LDL | mmol/l | 18 | Intercept & splines; unstructured | Intercept |
| **Medium LDL** | Concentration of medium LDL particles | mol/l | 18 | Intercept & splines; unstructured | Intercept |
|  | Total lipids in medium LDL | mmol/l | 18 | Intercept & splines; unstructured | Intercept |
|  | Phospholipids in medium LDL | mmol/l | 15 | Intercept & splines; matrix a = (1, 1, 0, 1, 1, 1) | Intercept |
|  | Total cholesterol in medium LDL | mmol/l | 18 | Intercept & splines; unstructured | Intercept |
|  | Cholesterol esters in medium LDL | mmol/l | 18 | Intercept & splines; unstructured | Intercept |
|  | Free cholesterol in medium LDL | mmol/l | 18 | Intercept & splines; unstructured | Intercept |
|  | Triglycerides in medium LDL | mmol/l | 18 | Intercept & splines; matrix a = (1, 1, 1, 1, 1, 0) | Intercept |
| **Small LDL** | Concentration of small LDL particles | mol/l | 18 | Intercept & splines; unstructured | Intercept |
|  | Total lipids in small LDL | mmol/l | 18 | Intercept & splines; unstructured | Intercept |
|  | Phospholipids in small LDL | mmol/l | 15 | Intercept & splines; matrix a = (1, 1, 0, 1, 1, 1) | Intercept |
|  | Total cholesterol in small LDL | mmol/l | 18 | Intercept & splines; unstructured | Intercept |
|  | Cholesterol esters in small LDL | mmol/l | 18 | Intercept & splines; unstructured | Intercept |
|  | Free cholesterol in small LDL | mmol/l | 18 | Intercept & splines; unstructured | Intercept |
|  | Triglycerides in small LDL | mmol/l | 15 | Intercept & splines; matrix a = (1, 1, 1, 1, 1, 0) | Intercept |
| **Very large HDL** | Concentration of very large HDL particles | mol/l | 15 | Intercept & splines; unstructured | Intercept |
|  | Total lipids in very large HDL | mmol/l | 15 | Intercept & splines; unstructured | Intercept |
|  | Phospholipids in very large HDL | mmol/l | 15 | Intercept & splines; unstructured | Intercept |
|  | Total cholesterol in very large HDL | mmol/l | 18 | Intercept & splines; unstructured | Intercept |
|  | Cholesterol esters in very large HDL | mmol/l | 18 | Intercept & splines; unstructured | Intercept |
|  | Free cholesterol in very large HDL | mmol/l | 15 | Intercept & splines; unstructured | Intercept |
|  | Triglycerides in very large HDL | mmol/l | 15 | Intercept & splines; unstructured | Intercept |
| **Large HDL** | Concentration of large HDL particles | mol/l | 18 | Intercept & splines; unstructured | Intercept |
|  | Total lipids in large HDL | mmol/l | 18 | Intercept & splines; unstructured | Intercept |
|  | Phospholipids in large HDL | mmol/l | 18 | Intercept & splines; unstructured | Intercept |
|  | Total cholesterol in large HDL | mmol/l | 18 | Intercept & splines; unstructured | Intercept |
|  | Cholesterol esters in large HDL | mmol/l | 18 | Intercept & splines; unstructured | Intercept |
|  | Free cholesterol in large HDL | mmol/l | 18 | Intercept & splines; unstructured | Intercept |
|  | Triglycerides in large HDL | mmol/l | 18 | Intercept & splines; unstructured | Intercept |
| **Medium HDL** | Concentration of medium HDL particles | mol/l | 18 | Intercept & splines; unstructured | Intercept |
|  | Total lipids in medium HDL | mmol/l | 15 | Intercept & splines; matrix a = (1, 1, 0, 1, 1, 1) | Intercept |
|  | Phospholipids in medium HDL | mmol/l | 15 | Intercept & splines; matrix a = (1, 1, 0, 1, 1, 1) | Intercept |
|  | Total cholesterol in medium HDL | mmol/l | 18 | Intercept & splines; unstructured | Intercept |
|  | Cholesterol esters in medium HDL | mmol/l | 18 | Intercept & splines; unstructured | Intercept |
|  | Free cholesterol in medium HDL | mmol/l | 18 | Intercept & splines; unstructured | Intercept |
|  | Triglycerides in medium HDL | mmol/l | 15 | Intercept & splines; matrix a = (1, 1, 0, 1, 1, 1) | Intercept |
| **Small HDL** | Concentration of small HDL particles | mol/l | 15 | Intercept & splines; matrix a = (0, 1, 0, 1, 1, 1) | Intercept |
|  | Total lipids in small HDL | mmol/l | 15 | Intercept & splines; matrix a = (1, 1, 0, 1, 1, 0) | Intercept |
|  | Phospholipids in small HDL | mmol/l | 15 | Intercept & splines; matrix a = (1, 1, 0, 1, 1, 1) | Intercept |
|  | Total cholesterol in small HDL | mmol/l | 15 | Intercept & splines; matrix a = (1, 1, 0, 1, 1, 1) | Intercept |
|  | Cholesterol esters in small HDL | mmol/l | 15 | Intercept & splines; matrix a = (1, 1, 0, 1, 1, 1) | Intercept |
|  | Free cholesterol in small HDL | mmol/l | 18 | Intercept & splines; unstructured | Intercept |
|  | Triglycerides in small HDL | mmol/l | 15 | Intercept & splines; unstructured | Intercept |
| **Lipoprotein particle size** | Mean diameter for VLDL particles | nm | 15 | Intercept & splines; unstructured | Intercept |
|  | Mean diameter for LDL particles | nm | 15 | Intercept & splines; matrix a = (1, 1, 0, 1, 1, 1) | Intercept |
|  | Mean diameter for HDL particles | nm | 15 | Intercept & splines; unstructured | Intercept |
| **Cholesterol concentrations** | Total cholesterol | mmol/l | 18 | Intercept & splines; unstructured | Intercept |
|  | Total cholesterol in VLDL | mmol/l | 18 | Intercept & splines; unstructured | Intercept |
|  | Remnant cholesterol (non-HDL and non-LDL cholesterol) | mmol/l | 18 | Intercept & splines; unstructured | Intercept |
|  | Total cholesterol in LDL | mmol/l | 18 | Intercept & splines; unstructured | Intercept |
|  | Total cholesterol in HDL | mmol/l | 18 | Intercept & splines; unstructured | Intercept |
|  | Total cholesterol in HDL2 | mmol/l | 18 | Intercept & splines; unstructured | Intercept |
|  | Total cholesterol in HDL3 | mmol/l | 18 | Intercept & splines; unstructured | Intercept |
|  | Esterified cholesterol | mmol/l | 15 | Intercept & splines; matrix a = (1, 1, 0, 1, 1, 1) | Intercept |
|  | Free cholesterol | mmol/l | 18 | Intercept & splines; unstructured | Intercept |
| **Glycerides and phospholipid concentrations** | Total triglycerides | mmol/l | 15 | Intercept & splines; unstructured | Intercept |
|  | Triglycerides in VLDL | mmol/l | 15 | Intercept & splines; unstructured | Intercept |
|  | Triglycerides in LDL | mmol/l | 18 | Intercept & splines; matrix a = (1, 1, 1, 1, 1, 0) | Intercept |
|  | Triglycerides in HDL | mmol/l | 15 | Intercept & splines; unstructured | Intercept |
|  | Diacylglycerol* | mmol/l | 15 | Intercept & splines; matrix a = (1, 1, 1, 1, 1, 0) | Intercept |
|  | Total phosphoglycerides (mmol/l) | mmol/l | 18 | Intercept & splines; matrix a = (1, 1, 0, 1, 1, 1) | Intercept |
|  | Phosphatidylcholine and other cholines (mmol/l) | mmol/l | 18 | Intercept & splines; unstructured | Intercept |
|  | Total cholines (mmol/l) | mmol/l | 15 | Intercept & splines; unstructured | Intercept |
| **Apolipoprotein concentrations** | Apolipoprotein A-1 | g/l | 18 | Intercept & splines; unstructured | Intercept |
|  | Apolipoprotein B | g/l | 15 | Intercept & splines; unstructured | Intercept |
| **Fatty acid concentrations** | Total fatty acids | mmol/l | 15 | Intercept & splines; unstructured | Intercept |
|  | Fatty acid length* |  | 15 | Intercept & splines; unstructured | Intercept |
|  | Estimated degree of unsaturation* |  | 15 | Intercept & splines; unstructured | Intercept |
|  | 22:6, docosahexaenoic acid | mmol/l | 15 | Intercept & splines; matrix a = (1, 1, 0, 1, 1, 1) | Intercept |
|  | 18:2 linoleic acid | mmol/l | 15 | Intercept & splines; unstructured | Intercept |
|  | Conjugated linoleic acid* | mmol/l | 15 | Intercept & splines; unstructured | Intercept |
|  | Omega-3 fatty acids | mmol/l | 15 | Intercept & splines; matrix a = (1, 1, 0, 1, 1, 1) | Intercept |
|  | Omega-6 fatty acids | mmol/l | 15 | Intercept & splines; unstructured | Intercept |
|  | Polyunsaturated fatty acids | mmol/l | 15 | Intercept & splines; unstructured | Intercept |
|  | Monounsaturated fatty acids; 16:1, 18:1 | mmol/l | 18 | Intercept & splines; unstructured | Intercept |
|  | Saturated fatty acids | mmol/l | 15 | Intercept & splines; unstructured | Intercept |
| **Glycolysis related metabolite** | Glucose | mmol/l | 15 | Intercept & splines; matrix a = (1, 1, 1, 1, 1, 0) | Intercept |
|  | Lactate | mmol/l | 15 | Intercept & splines; unstructured | Intercept |
|  | Citrate | mmol/l | 18 | Intercept & splines; unstructured | Intercept |
| **Amino acid concentrations** | Alanine | mmol/l | 18 | Intercept & splines; unstructured | Intercept |
|  | Glutamine | mmol/l | 18 | Intercept & splines; unstructured | Intercept |
|  | Histidine | mmol/l | 18 | Intercept & splines; matrix a = (1, 1, 1, 1, 1, 0) | Intercept |
| branched | Isoleucine | mmol/l | 18 | Intercept & splines; matrix a = (1, 1, 1, 1, 1, 0) | Intercept |
| branched | Leucine | mmol/l | 18 | Intercept & splines; unstructured | Intercept |
| branched | Valine | mmol/l | 15 | Intercept & splines; unstructured | Intercept |
| aromatic | Phenylalanine | mmol/l | 18 | Intercept & splines; unstructured | Intercept |
| aromatic | Tyrosine | mmol/l | 15 | Intercept & splines; matrix a = (1, 1, 1, 1, 1, 0) | Intercept |
| **Ketone body concentrations** | Acetate | mmol/l | 15 | Intercept & splines; matrix a = (1, 1, 0, 1, 1, 1) | Intercept |
|  | Acetoacetate | mmol/l | Linear | Intercept & slope; unstructured | Intercept |
|  | 3-hydroxybutyrate | mmol/l | 18 | Intercept & splines; matrix a = (0, 1, 1, 1, 1, 1) | Intercept |
| **Fluid balance marker** | Creatinine | mmol/l | 18 | Intercept & splines; unstructured | Intercept |
|  | Albumin | mmol/l | 18 | Intercept & splines; matrix a = (1, 1, 0, 1, 1, 1) | Intercept |
| **Inflammation marker** | Glycoprotein acetyls, mainly a1-acid glycoprotein | mmol/l | 18 | Intercept & splines; unstructured | Intercept |
| *These metabolites were not measured at 25y; all models include data only up to aged 18y. HDL: high-density lipoprotein; IDL: intermediate-density lipoprotein; LDL: low-density lipoprotein; VLDL: very-low-density lipoprotein. | | | | | |

**eMethods 3**

**Characteristics of included vs. excluded participants**

We examined characteristics associated with not being included in our analyses due to missing cardiometabolic trait data or attrition from the cohort. To do this, we compared the socio-demographic characteristics at birth of mothers and partners of participants included in the main analyses compared to those excluded from the analyses. All characteristics were measured during pregnancy or at birth through questionnaires or from routine health records.

Marital status was obtained from antenatal questionnaires and classified as never married, widowed, divorced, separated, first marriage, marriage 2 or 3. Smoking in the first trimester of pregnancy was self-reported by mothers at 18 weeks gestation. Birthweight and gestational age were derived from clinical records. Maternal age was reported in the mother’s antenatal questionnaires. Maternal pre-pregnancy weight and height were self-reported in antenatal questionnaires.

We performed weighted sensitivity analyses using inverse probability weighting to address potential selection bias. The participant level weights were estimated using logistic regression using all socio-demographic characteristics listed above with the addition of gender and were subsequently incorporated into the multi-level models as level two weights which adjust for the unequal probability of selection of the participants.

|  | **eTable 3: Characteristics of offspring included in analyses compared to those excluded due to missing cardiometabolic trait data** | | | | | |  |
| --- | --- | --- | --- | --- | --- | --- | --- |
|  | | **Female participants included**  **n=3,300*** | **Female participants excluded** | **N excluded females** | **Male participants included**  **n=3,237*** | **Male participants excluded** | **N excluded males** |
|  | | **n (%)** | **n (%)** | **n** | **n (%)** | **n (%)** | **N** |
| **Non-white ethnicity** | | 68 (2.1) | 84 (3.1) | 2,708 | 56 (1.7) | 116 (3.7) | 3,168 |
| **Maternal marital status** | |  |  | 3,122 |  |  | 3,642 |
| Never married | | 464 (14.2) | 743 (23.8) |  | 419 (13.1) | 901 (24.7) |  |
| Separated/Divorced/Widowed | | 147 (4.5) | 222 (7.1) |  | 148 (4.6) | 274 (7.5) |  |
| 1^st^ Marriage | | 2442 (74.9) | 1971 (63.1) |  | 2407 (75.4) | 2223 (61.0) |  |
| Marriage 2 or 3 | | 206 (6.3) | 186 (6.0) |  | 219 (6.9) | 244 (6.7) |  |
| **Household social class** † | |  |  | 2,460 |  |  | 2,865 |
| Professional | | 502 (15.9) | 228 (9.3) |  | 519 (16.9) | 288 (10.1) |  |
| Managerial & Technical | | 1409 (44.7) | 947 (38.5) |  | 1356 (44.1) | 1110 (38.7) |  |
| Non-Manual | | 765 (24.3) | 665 (27.0) |  | 765 (24.9) | 750 (26.2) |  |
| Manual | | 329 (10.4) | 425 (17.3) |  | 317 (10.3) | 490 (17.1) |  |
| Part Skilled & Unskilled | | 149 (4.7) | 195 (7.9) |  | 120 (3.9) | 227 (7.9) |  |
| **Maternal education** | |  |  | 2,727 |  |  | 3,201 |
| Less than O level | | 725 (22.0) | 1061 (38.9) |  | 692 (21.4) | 1270 (39.7) |  |
| O level | | 1125 (34.1) | 961 (35.2) |  | 1158 (35.8) | 1073 (33.5) |  |
| A level | | 890 (27.0) | 471 (17.3) |  | 855 (26.4) | 578 (18.1) |  |
| Degree or above | | 560 (17.0) | 234 (8.6) |  | 532 (16.4) | 280 (8.7) |  |
| **Mother’s Partner’s highest educational qualification** | |  |  | 2,605 |  |  | 3,025 |
| Less than O level | | 917 (28.6) | 1104 (42.4) |  | 825 (26.2) | 1296 (42.8) |  |
| O level | | 690 (21.5) | 528 (20.3) |  | 697 (22.2) | 636 (21.0) |  |
| A level | | 884 (27.5) | 657 (25.2) |  | 889 (28.3) | 686 (22.7) |  |
| Degree or Above | | 718 (22.4) | 316 (12.1) |  | 734 (23.3) | 407 (13.5) |  |
| **Maternal smoking during pregnancy** | | 603 (21.6) | 793 (29.3) | 2,711 | 825 (26.2) | 1296 (42.8) | 3,119 |
|  | | ***Mean (SD)*** | ***Mean (SD)*** |  | ***Mean (SD)*** | ***Mean (SD)*** |  |
| **Birthweight (g)** | | 3375 (504.0) | 3285 (580.2) | 3,444 | 3478 (572.5) | 3392 (631.2) | 3,978 |
| **Gestational age (weeks)** | | 40 (1.7) | 39 (2.8) | 3,522 | 39 (1.9) | 39 (2.9) | 4,071 |
| **Maternal age (years)** | | 29 (4.5) | 27 (4.9) | 2,551 | 29 (4.5) | 27 (5.0) | 3,039 |
| **Maternal pre-pregnancy BMI (kg/m^2^)** | | 23 (3.4) | 23(3.6) | 2,616 | 23 (3.4) | 23(3.6) | 2,999 |

^*^Represents participants included in models of 144 concentrations; exact denominators in this table will vary due to missing data for characteristics which were not required for inclusion in analyses. Denominators for excluded participants also vary.

**eTable 4:** Number of participants with measures at each time point*

|  | **Age 7** | **Age 15** | **Age 18** | **Age 25** |
| --- | --- | --- | --- | --- |
| **Females: n (%)** |  |  |  |  |
| Less than O-level | 467 (37.7) | 249 (20.1) | 256 (20.7) | 266 (21.5) |
| O-level | 728 (33.4) | 477 (21.9) | 456 (20.9) | 518 (23.8) |
| A-level | 609 (33.7) | 369 (20.4) | 377 (20.8) | 452 (25.0) |
| Degree or above | 390 (31.8) | 254 (20.7) | 270 (22.0) | 312 (25.4) |
| **Males: n (%)** |  |  |  |  |
| Less than O-level | 509 (45.7) | 217 (19.5) | 223 (20.0) | 165 (14.8) |
| O-level | 892 (44.0) | 410 (20.3) | 391 (19.3) | 332 (16.4) |
| A-level | 615 (35.9) | 386 (22.6) | 401 (23.4) | 309 (18.1) |
| Degree or above | 395 (34.3) | 246 (21.3) | 266 (23.1) | 246 (21.3) |

*Represents participants included in models of 144 concentrations

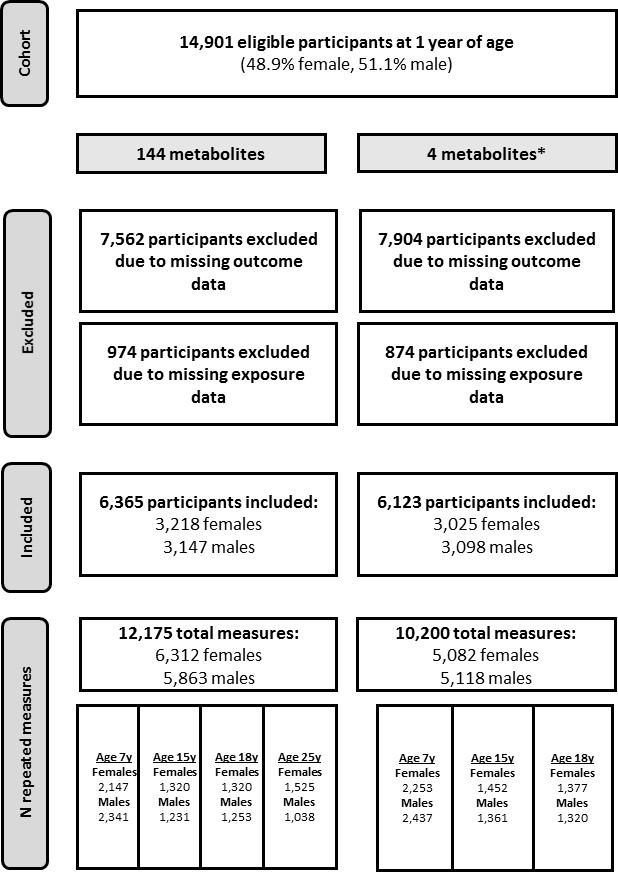

**eFigure 1: Flowchart of participants included in paternal education analyses**

*Four metabolites (diacylglycerol, fatty acid chain length, estimated degree of unsaturation and conjugated linoleic acid) were not measured at 25y; thus, for these traits change over time is only modelled to age 18y.

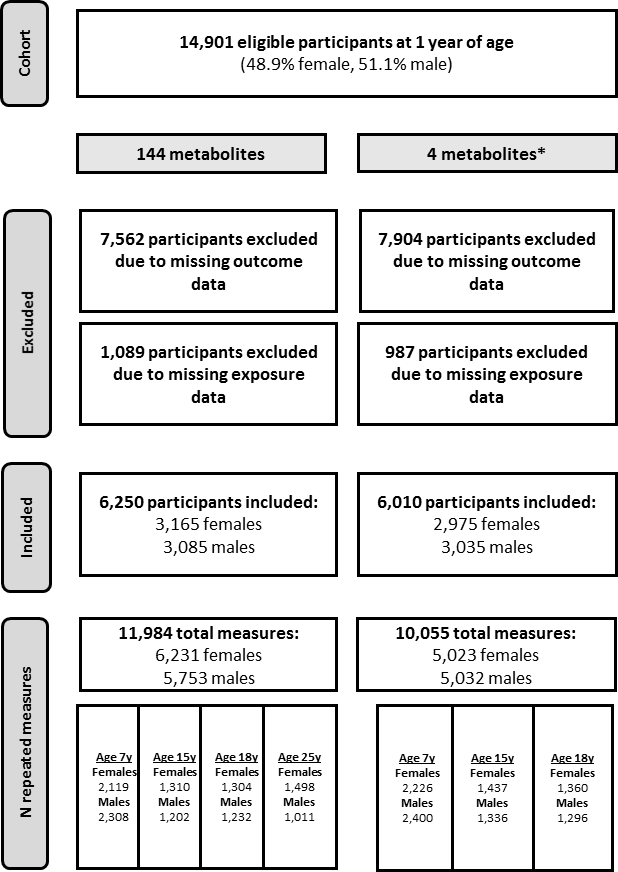

**eFigure 2: Flowchart of participants included in household social class analyses**

*Four metabolites (diacylglycerol, fatty acid chain length, estimated degree of unsaturation and conjugated linoleic acid) were not measured at 25y; thus, for these traits change over time is only modelled to age 18y.

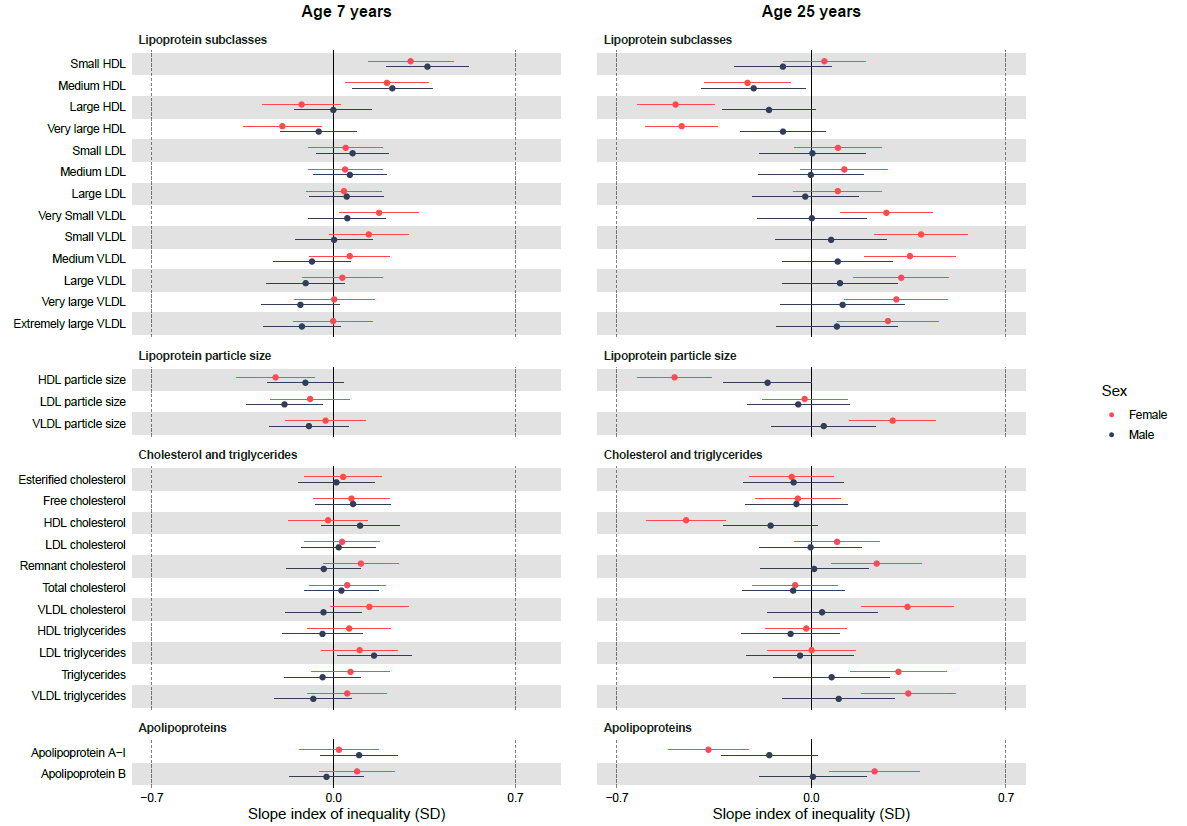

**eFigure 3: Sex-specific associations of paternal education with lipoprotein and lipid concentrations at 7y and 25y, estimated from multilevel models**

Slope index of inequality represents the mean difference in SDs of the outcome between the individuals of lowest and highest socioeconomic position on the hypothetical underlying continuous distribution of paternal education. Results shown are standardised mean differences in metabolic traits comparing lowest paternal education (less than O-level) with highest paternal education (degree level) with whiskers representing 95% confidence intervals. HDL, high-density lipoprotein; LDL, low-density lipoprotein; SD, standard deviation; VLDL, very-low-density lipoprotein.

|  | **Females** | | **Males** | |
| --- | --- | --- | --- | --- |
|  | **Mean change (95% CI) in concentration in degree level maternal education** | **Mean difference (95% CI) in change in less than O-level maternal education** | **Mean change (95% CI) in concentration in degree level maternal education** | **Mean difference (95% CI) in change in less than O-level maternal education** |
| **Lipoprotein subclasses** |  |  |  |  |
| Small HDL | 3.40 (3.19,3.60) | -0.18 (-0.55,0.18) | 3.01 (2.78,3.24) | -0.61 (-1.04,-0.17) |
| Medium HDL | 4.32 (4.09,4.55) | -0.79 (-1.21,-0.38) | 2.57 (2.31,2.83) | -0.76 (-1.25,-0.26) |
| Large HDL | 1.65 (1.47,1.82) | -0.97 (-1.28,-0.66) | -0.76 (-0.96,-0.57) | -0.34 (-0.71,0.03) |
| Very large HDL | -0.11 (-0.24,0.02) | -0.61 (-0.85,-0.38) | -1.88 (-2.02,-1.73) | -0.12 (-0.39,0.15) |
| Small LDL | 0.40 (0.26,0.54) | 0.10 (-0.15,0.36) | 0.63 (0.47,0.79) | -0.07 (-0.37,0.24) |
| Medium LDL | 0.80 (0.66,0.93) | 0.13 (-0.11,0.38) | 1.04 (0.88,1.19) | -0.07 (-0.35,0.22) |
| Large LDL | 0.82 (0.69,0.96) | 0.10 (-0.14,0.35) | 0.91 (0.76,1.06) | -0.08 (-0.37,0.21) |
| Very Small VLDL | -0.83 (-0.97,-0.69) | 0.23 (-0.03,0.48) | -0.48 (-0.64,-0.32) | -0.05 (-0.35,0.25) |
| Small VLDL | -1.03 (-1.16,-0.91) | 0.33 (0.11,0.56) | -0.16 (-0.30,-0.03) | 0.08 (-0.18,0.34) |
| Medium VLDL | -0.52 (-0.64,-0.41) | 0.31 (0.10,0.52) | 0.23 (0.10,0.36) | 0.18 (-0.05,0.42) |
| Large VLDL | -0.58 (-0.69,-0.47) | 0.24 (0.04,0.43) | -0.07 (-0.19,0.05) | 0.19 (-0.02,0.41) |
| Very large VLDL | -0.74 (-0.84,-0.64) | 0.17 (0.01,0.35) | -0.39 (-0.49,-0.29) | 0.19 (0.01,0.38) |
| Extremely large VLDL | -0.75 (-0.85,-0.66) | 0.15 (-0.02,0.32) | -0.42 (-0.52,-0.33) | 0.17 (-0.01,0.35) |
| **Lipoprotein particle size** |  |  |  |  |
| HDL particle size | 0.004 (-0.13,0.13) | -0.60 (-0.83,-0.37) | -1.75 (-1.89,-1.60) | -0.16 (-0.42,0.11) |
| LDL particle size | -0.09 (-0.23,0.04) | 0.06 (-0.19,0.30) | -0.64 (-0.80,-0.49) | 0.12 (-0.16,0.40) |
| VLDL particle size | -0.54 (-0.65,-0.44) | 0.28 (0.09,0.47) | 0.16 (0.04,0.27) | 0.13 (-0.07,0.34) |
| **Cholesterol and triglycerides** | |  |  |  |
| Esterified cholesterol | -0.58 (-0.71,-0.46) | -0.13 (-0.35,0.09) | -0.84 (-0.97,-0.70) | -0.10 (-0.35,0.16) |
| Free cholesterol | -0.50 (-0.62,-0.38) | -0.13 (-0.35,0.08) | -0.76 (-0.89,-0.63) | -0.14 (-0.39,0.11) |
| HDL cholesterol | 0.40 (0.26,0.54) | -0.73 (-0.98,-0.48) | -1.27 (-1.43,-1.12) | -0.35 (-0.64,-0.05) |
| LDL cholesterol | -0.23 (-0.35,-0.12) | 0.08 (-0.13,0.29) | 0.03 (-0.10,0.16) | -0.02 (-0.27,0.22) |
| Remnant cholesterol | -1.46 (-1.58,-1.34) | 0.18 (-0.03,0.39) | -0.99 (-1.12,-0.85) | 0.05 (-0.20,0.30) |
| Total cholesterol | -0.57 (-0.69,-0.45) | -0.13 (-0.35,0.09) | -0.80 (-0.94,-0.67) | -0.12 (-0.38,0.14) |
| VLDL cholesterol | -1.66 (-1.77,-1.54) | 0.25 (0.04,0.45) | -0.98 (-1.11,-0.86) | 0.08 (-0.16,0.32) |
| HDL triglycerides | 0.48 (0.34,0.61) | -0.08 (-0.32,0.15) | -0.13 (-0.27,0.02) | -0.06 (-0.33,0.21) |
| LDL triglycerides | -0.25 (-0.36,-0.15) | -0.10 (-0.29,0.09) | -0.61 (-0.72,-0.50) | -0.19 (-0.39,0.01) |
| Triglycerides | -0.68 (-0.79,-0.57) | 0.21 (0.01,0.41) | -0.23 (-0.35,-0.11) | 0.11 (-0.11,0.33) |
| VLDL triglycerides | -0.74 (-0.85,-0.63) | 0.26 (0.06,0.46) | -0.11 (-0.22,0.01) | 0.17 (-0.05,0.38) |
| **Apolipoproteins** |  |  |  |  |
| Apolipoprotein A-I | 0.51 (0.35,0.66) | -0.71 (-0.99,-0.43) | -0.99 (-1.16,-0.81) | -0.38 (-0.71,-0.05) |
| Apolipoprotein B | -0.77 (-0.89,-0.65) | 0.19 (-0.03,0.40) | -0.20 (-0.33,-0.07) | 0.03 (-0.21,0.28) |

**eTable 8: Mean change in lipoprotein and lipid concentrations between 7y and 25y, by paternal education, estimated from multilevel models**

Results shown are standardised mean change and mean difference in metabolic traits. The intercept, spline periods and coefficients for the slope index of inequality from multilevel models were used to estimate the mean change in concentration for degree level maternal education and mean difference for less than O-level. CI, confidence interval; HDL, high-density lipoprotein; LDL, low-density lipoprotein; VLDL, very-low-density lipoprotein.

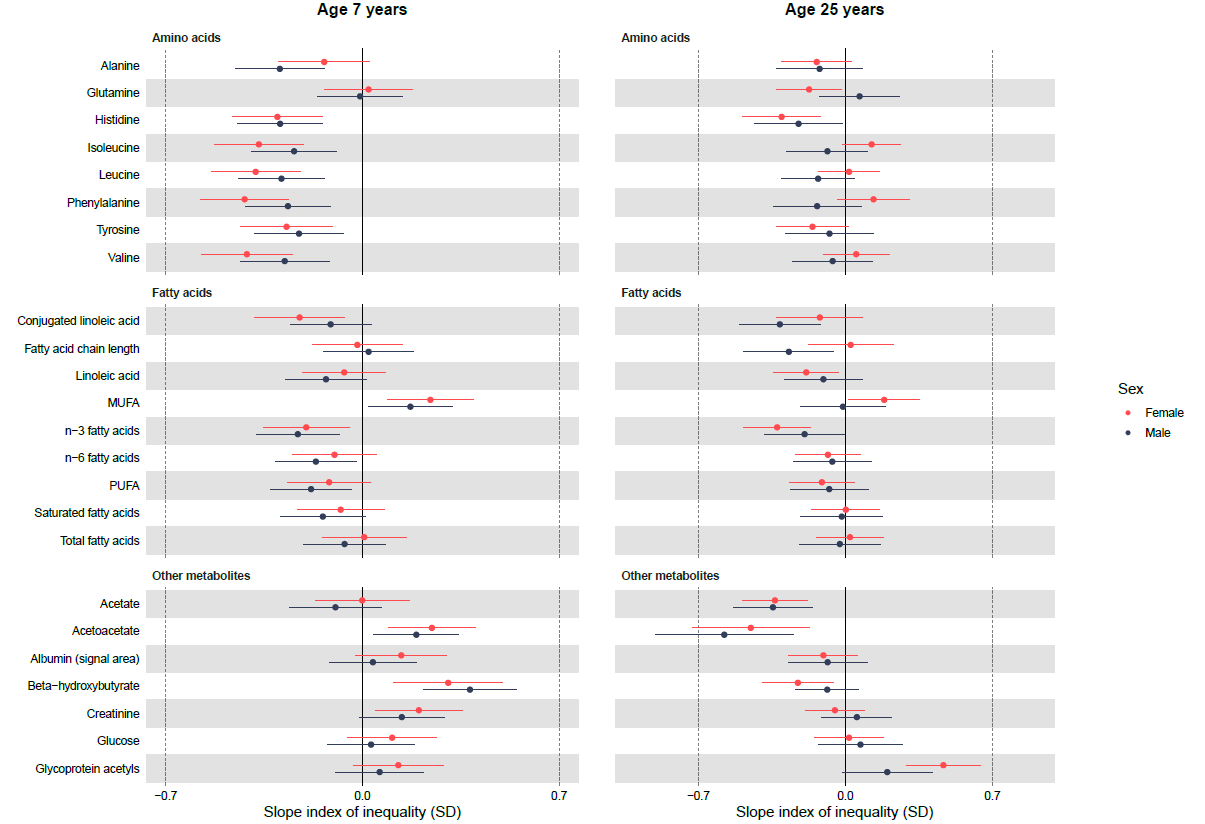

**eFigure 4: Sex-specific associations of paternal education with amino acids, fatty acids and other metabolic at 7y and 25y, estimated from multilevel models**

Slope index of inequality represents the mean difference in SDs of the outcome between the individuals of lowest and highest socioeconomic position on the hypothetical underlying continuous distribution of maternal education. Results shown are standardised mean differences in metabolic traits comparing lowest paternal education (less than O-level) with highest paternal education (degree level) with whiskers representing 95% confidence intervals. MUFA, monounsaturated fatty acids; PUFA, polyunsaturated fatty acids; SD, standard deviation; VLDL, very-low-density lipoprotein. *Conjugated linoleic acid and fatty acid chain length are modelled up to 18y only.

|  | **Females** | | **Males** | |
| --- | --- | --- | --- | --- |
|  | **Mean change (95% CI) in concentration in degree level maternal education** | **Mean difference (95% CI) in change in less than O-level maternal education** | **Mean change (95% CI) in concentration in degree level maternal education** | **Mean difference (95% CI) in change in less than O-level maternal education** |
| **Amino acids** |  |  |  |  |
| Alanine | 0.63 (0.51,0.75) | 0.02 (-0.20,0.23) | 0.78 (0.65,0.90) | 0.19 (-0.05,0.42) |
| Glutamine | -2.33 (-2.47,-2.20) | -0.24 (-0.49,0.0002) | -1.02 (-1.17,-0.87) | 0.10 (-0.18,0.38) |
| Histidine | -2.22 (-2.33,-2.11) | 0.11 (-0.09,0.30) | -1.81 (-1.92,-1.70) | 0.15 (-0.05,0.35) |
| Isoleucine | -0.72 (-0.82,-0.62) | 0.44 (0.27,0.62) | 0.03 (-0.08,0.13) | 0.19 (0.004,0.38) |
| Leucine | -0.38 (-0.49,-0.27) | 0.39 (0.20,0.59) | 0.49 (0.38,0.61) | 0.18 (-0.02,0.39) |
| Phenylalanine | 0.28 (0.16,0.39) | 0.52 (0.32,0.72) | 0.67 (0.55,0.78) | 0.16 (-0.05,0.38) |
| Tyrosine | -1.82 (-1.93,-1.71) | 0.18 (-0.01,0.37) | -1.46 (-1.57,-1.35) | 0.18 (-0.02,0.38) |
| Valine | -0.57 (-0.68,-0.45) | 0.45 (0.26,0.65) | 0.30 (0.19,0.42) | 0.23 (0.02,0.44) |
| **Fatty acids** |  |  |  |  |
| Conjugated linoleic acid | -0.58 (-0.71,-0.46) | 0.14 (-0.07,0.35) | -0.45 (-0.56,-0.34) | -0.11 (-0.29,0.08) |
| Fatty acid chain length | 0.46 (0.31,0.60) | 0.04 (-0.21,0.30) | 0.64 (0.49,0.79) | -0.30 (-0.56,-0.04) |
| Linoleic acid | -1.10 (-1.22,-0.98) | -0.16 (-0.37,0.06) | -1.28 (-1.41,-1.15) | 0.01 (-0.24,0.25) |
| MUFA | -0.46 (-0.59,-0.33) | -0.03 (-0.26,0.20) | -0.40 (-0.54,-0.25) | -0.18 (-0.44,0.08) |
| n-3 fatty acids | -0.53 (-0.65,-0.40) | -0.17 (-0.39,0.05) | -0.58 (-0.71,-0.45) | 0.01 (-0.24,0.26) |
| n-6 fatty acids | -1.13 (-1.25,-1.01) | -0.001 (-0.22,0.22) | -1.40 (-1.53,-1.27) | 0.09 (-0.16,0.34) |
| PUFA | -1.11 (-1.24,-0.99) | -0.02 (-0.24,0.20) | -1.37 (-1.50,-1.23) | 0.09 (-0.16,0.34) |
| Saturated fatty acids | -1.41 (-1.53,-1.28) | 0.08 (-0.14,0.30) | -1.58 (-1.71,-1.44) | 0.12 (-0.12,0.37) |
| Total fatty acids | -1.11 (-1.23,-0.98) | 0.02 (-0.20,0.24) | -1.25 (-1.38,-1.11) | 0.03 (-0.22,0.29) |
| **Other metabolites** |  |  |  |  |
| Acetate | -0.49 (-0.59,-0.38) | -0.18 (-0.36,0.01) | -0.39 (-0.50,-0.29) | -0.09 (-0.28,0.10) |
| Acetoacetate | -0.45 (-0.56,-0.34) | -0.38 (-0.57,-0.19) | -0.39 (-0.50,-0.28) | -0.36 (-0.56,-0.16) |
| Albumin | 0.22 (0.06,0.39) | -0.30 (-0.59,-0.01) | 1.28 (1.11,1.46) | -0.17 (-0.50,0.16) |
| Beta-hydroxybutyrate | 1.33 (1.17,1.50) | -0.58 (-0.86,-0.30) | 1.18 (1.05,1.31) | -0.49 (-0.73,-0.24) |
| Creatinine | 3.08 (2.95,3.22) | -0.28 (-0.52,-0.03) | 5.10 (4.95,5.25) | -0.06 (-0.33,0.22) |
| Glucose | -0.52 (-0.63,-0.42) | -0.09 (-0.28,0.09) | -0.38 (-0.48,-0.27) | 0.01 (-0.18,0.20) |
| Glycoprotein acetyls | -0.28 (-0.42,-0.14) | 0.42 (0.17,0.67) | -0.13 (-0.29,0.02) | 0.17 (-0.11,0.46) |

**eTable 9:** **Mean change in amino acids, fatty acids and other metabolic concentrations between 7y and 25y in females, by paternal education, estimated from multilevel models**

Results shown are standardised mean change and mean difference in metabolic traits. The intercept, spline periods and coefficients for the slope index of inequality from multilevel models were used to estimate the mean change in concentration for degree level maternal education and mean difference for less than O-level. CI, confidence interval; MUFA, monounsaturated fatty acids; PUFA, polyunsaturated fatty acids; VLDL, very-low-density lipoprotein. *Conjugated linoleic acid and fatty acid chain length are modelled up to 18y only.

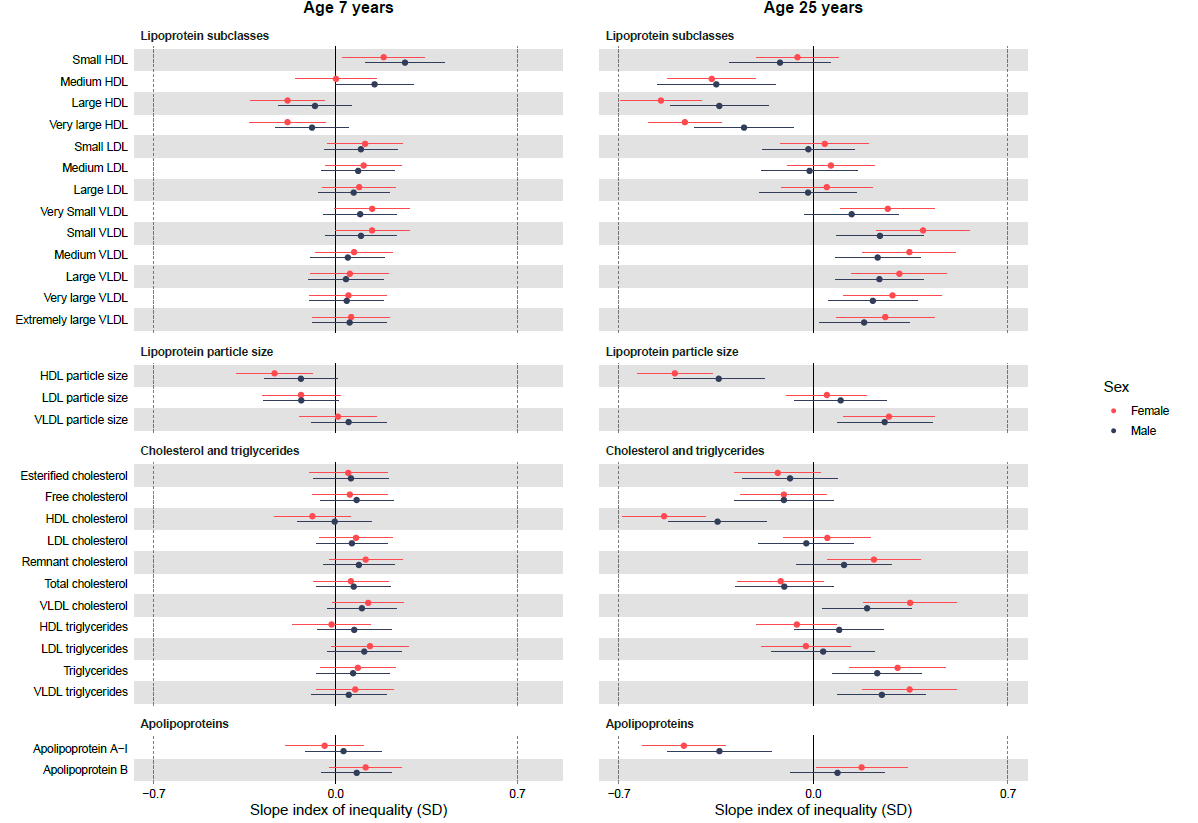

**eFigure 5: Sex-specific associations of household social class with lipoprotein and lipid concentrations at 7y and 25y, estimated from multilevel models**

Slope index of inequality represents the mean difference in SDs of the outcome between the individuals of lowest and highest socioeconomic position on the hypothetical underlying continuous distribution of household social class. Results shown are standardised mean differences in metabolic traits comparing lowest household social class with highest household social class with whiskers representing 95% confidence intervals. HDL, high-density lipoprotein; LDL, low-density lipoprotein; SD, standard deviation; VLDL, very-low-density lipoprotein.

|  | **Females** | | **Males** | |
| --- | --- | --- | --- | --- |
|  | **Mean change (95% CI) in concentration in degree level maternal education** | **Mean difference (95% CI) in change in less than O-level maternal education** | **Mean change (95% CI) in concentration in degree level maternal education** | **Mean difference (95% CI) in change in less than O-level maternal education** |
| **Lipoprotein subclasses** |  |  |  |  |
| Small HDL | 3.32 (3.15,3.49) | -0.32 (-0.69,0.04) | 2.83 (2.64,3.02) | -0.55 (-0.98,-0.12) |
| Medium HDL | 4.29 (4.10,4.48) | -0.94 (-1.37,-0.52) | 2.55 (2.33,2.77) | -0.97 (-1.48,-0.46) |
| Large HDL | 1.52 (1.38,1.66) | -0.98 (-1.29,-0.67) | -0.68 (-0.84,-0.52) | -0.63 (-1.00,-0.26) |
| Very large HDL | -0.16 (-0.27,-0.05) | -0.62 (-0.85,-0.38) | -1.81 (-1.93,-1.69) | -0.29 (-0.56,-0.01) |
| Small LDL | 0.49 (0.37,0.62) | -0.05 (-0.32,0.23) | 0.64 (0.50,0.79) | -0.13 (-0.46,0.19) |
| Medium LDL | 0.90 (0.78,1.02) | -0.01 (-0.27,0.25) | 1.07 (0.93,1.20) | -0.11 (-0.42,0.20) |
| Large LDL | 0.87 (0.75,0.98) | -0.02 (-0.27,0.23) | 0.87 (0.74,1.00) | -0.10 (-0.40,0.19) |
| Very Small VLDL | -0.79 (-0.91,-0.67) | 0.25 (-0.004,0.51) | -0.54 (-0.67,-0.41) | 0.15 (-0.15,0.45) |
| Small VLDL | -0.94 (-1.04,-0.84) | 0.32 (0.10,0.54) | -0.20 (-0.31,-0.09) | 0.26 (0.01,0.51) |
| Medium VLDL | -0.47 (-0.57,-0.38) | 0.29 (0.08,0.50) | 0.23 (0.12,0.33) | 0.27 (0.03,0.51) |
| Large VLDL | -0.54 (-0.63,-0.45) | 0.21 (0.01,0.41) | -0.05 (-0.15,0.05) | 0.23 (0.01,0.45) |
| Very large VLDL | -0.68 (-0.77,-0.60) | 0.12 (-0.05,0.30) | -0.33 (-0.42,-0.25) | 0.13 (-0.05,0.32) |
| Extremely large VLDL | -0.70 (-0.78,-0.62) | 0.08 (-0.08,0.25) | -0.36 (-0.44,-0.28) | 0.08 (-0.09,0.26) |
| **Lipoprotein particle size** |  |  |  |  |
| HDL particle size | -0.05 (-0.16,0.06) | -0.58 (-0.82,-0.35) | -1.64 (-1.75,-1.52) | -0.42 (-0.69,-0.15) |
| LDL particle size | -0.15 (-0.27,-0.03) | 0.21 (-0.04,0.46) | -0.67 (-0.80,-0.54) | 0.29 (0.001,0.58) |
| VLDL particle size | -0.48 (-0.57,-0.39) | 0.22 (0.03,0.40) | 0.16 (0.06,0.25) | 0.20 (-0.01,0.40) |
| **Cholesterol and triglycerides** | |  |  |  |
| Esterified cholesterol | -0.57 (-0.68,-0.47) | -0.23 (-0.46,0.002) | -0.84 (-0.96,-0.73) | -0.19 (-0.45,0.08) |
| Free cholesterol | -0.49 (-0.59,-0.39) | -0.20 (-0.42,0.03) | -0.76 (-0.87,-0.65) | -0.23 (-0.49,0.03) |
| HDL cholesterol | 0.34 (0.22,0.45) | -0.75 (-1.00,-0.51) | -1.20 (-1.33,-1.08) | -0.55 (-0.84,-0.26) |
| LDL cholesterol | -0.19 (-0.28,-0.09) | -0.02 (-0.23,0.20) | 0.04 (-0.06,0.15) | -0.10 (-0.35,0.15) |
| Remnant cholesterol | -1.39 (-1.49,-1.29) | 0.14 (-0.07,0.36) | -0.96 (-1.06,-0.85) | 0.07 (-0.18,0.31) |
| Total cholesterol | -0.55 (-0.66,-0.45) | -0.22 (-0.45,0.01) | -0.80 (-0.92,-0.68) | -0.23 (-0.50,0.04) |
| VLDL cholesterol | -1.52 (-1.61,-1.43) | 0.24 (0.04,0.44) | -0.92 (-1.02,-0.82) | 0.15 (-0.07,0.38) |
| HDL triglycerides | 0.45 (0.34,0.56) | -0.06 (-0.29,0.17) | -0.15 (-0.27,-0.04) | 0.06 (-0.20,0.33) |
| LDL triglycerides | -0.24 (-0.33,-0.15) | -0.16 (-0.35,0.04) | -0.70 (-0.79,-0.60) | -0.08 (-0.29,0.14) |
| Triglycerides | -0.64 (-0.73,-0.54) | 0.18 (-0.01,0.38) | -0.24 (-0.34,-0.14) | 0.20 (-0.02,0.42) |
| VLDL triglycerides | -0.70 (-0.79,-0.60) | 0.24 (0.04,0.44) | -0.10 (-0.20,-0.004) | 0.24 (0.02,0.46) |
| **Apolipoproteins** |  |  |  |  |
| Apolipoprotein A-I | 0.49 (0.36,0.62) | -0.82 (-1.11,-0.53) | -0.97 (-1.12,-0.81) | -0.64 (-0.99,-0.29) |
| Apolipoprotein B | -0.69 (-0.78,-0.59) | 0.09 (-0.12,0.31) | -0.19 (-0.30,-0.09) | 0.04 (-0.21,0.29) |

**eTable 10:** **Mean change in lipoprotein and lipid concentrations between 7y and 25y, by household social class, estimated from multilevel models**

Results shown are standardised mean change and mean difference in metabolic traits. The intercept, spline periods and coefficients for the slope index of inequality from multilevel models were used to estimate the mean change in concentration for degree level maternal education and mean difference for less than O-level. CI, confidence interval; HDL, high-density lipoprotein; LDL, low-density lipoprotein; VLDL, very-low-density lipoprotein.

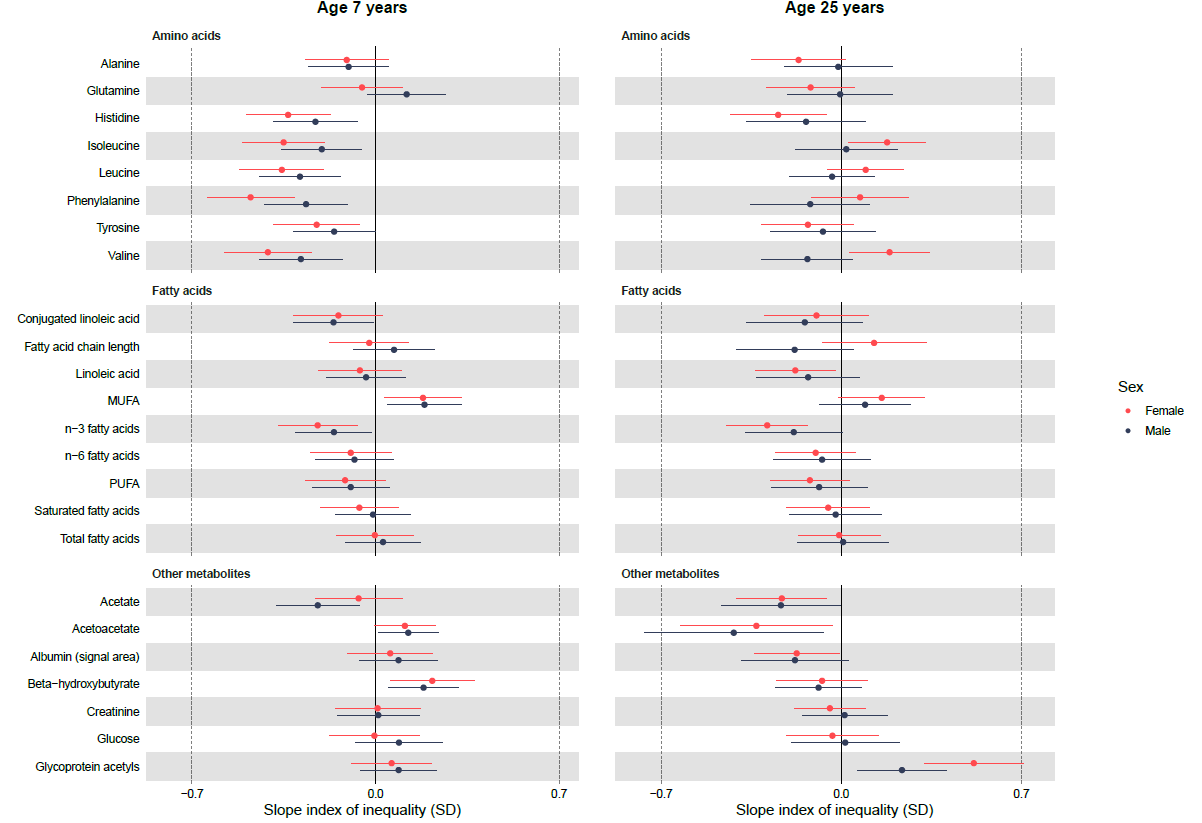

**eFigure 6: Sex-specific associations of household social class with amino acids, fatty acids and other metabolic at 7y and 25y, estimated from multilevel models**

Slope index of inequality represents the mean difference in SDs of the outcome between the individuals of lowest and highest socioeconomic position on the hypothetical underlying continuous distribution of household social class. Results shown are standardised mean differences in metabolic traits comparing lowest household social class with household social class with whiskers representing 95% confidence intervals. MUFA, monounsaturated fatty acids; PUFA, polyunsaturated fatty acids; SD, standard deviation; VLDL, very-low-density lipoprotein. *Conjugated linoleic acid and fatty acid chain length are modelled up to 18y only.

|  | **Females** | | **Males** | |
| --- | --- | --- | --- | --- |
|  | **Mean change (95% CI) in concentration in degree level maternal education** | **Mean difference (95% CI) in change in less than O-level maternal education** | **Mean change (95% CI) in concentration in degree level maternal education** | **Mean difference (95% CI) in change in less than O-level maternal education** |
| **Amino acids** |  |  |  |  |
| Alanine | 0.65 (0.55,0.75) | -0.03 (-0.24,0.19) | 0.83 (0.73,0.94) | 0.09 (-0.14,0.33) |
| Glutamine | -2.46 (-2.57,-2.34) | -0.10 (-0.35,0.16) | -0.94 (-1.06,-0.81) | -0.12 (-0.42,0.17) |
| Histidine | -2.38 (-2.48,-2.28) | 0.16 (-0.04,0.36) | -1.90 (-2.00,-1.80) | 0.13 (-0.09,0.36) |
| Isoleucine | -0.70 (-0.79,-0.61) | 0.47 (0.29,0.64) | 0.06 (-0.03,0.15) | 0.22 (0.01,0.43) |
| Leucine | -0.36 (-0.46,-0.27) | 0.44 (0.24,0.64) | 0.52 (0.42,0.62) | 0.26 (0.04,0.47) |
| Phenylalanine | 0.37 (0.27,0.47) | 0.53 (0.32,0.75) | 0.76 (0.65,0.86) | 0.17 (-0.06,0.41) |
| Tyrosine | -1.80 (-1.89,-1.71) | 0.15 (-0.05,0.34) | -1.41 (-1.51,-1.32) | 0.11 (-0.09,0.31) |
| Valine | -0.58 (-0.68,-0.48) | 0.57 (0.37,0.78) | 0.38 (0.28,0.48) | 0.16 (-0.06,0.38) |
| **Fatty acids** |  |  |  |  |
| Conjugated linoleic acid | -0.61 (-0.72,-0.50) | 0.07 (-0.15,0.29) | -0.56 (-0.66,-0.46) | 0.06 (-0.16,0.27) |
| Fatty acid chain length | 0.42 (0.30,0.53) | 0.15 (-0.09,0.39) | 0.56 (0.44,0.68) | -0.24 (-0.49,0.01) |
| Linoleic acid | -1.20 (-1.31,-1.09) | -0.18 (-0.41,0.06) | -1.34 (-1.46,-1.22) | -0.13 (-0.40,0.15) |
| MUFA | -0.45 (-0.56,-0.35) | 0.001 (-0.23,0.23) | -0.45 (-0.56,-0.33) | -0.06 (-0.33,0.20) |
| n-3 fatty acids | -0.56 (-0.67,-0.46) | -0.13 (-0.35,0.10) | -0.57 (-0.69,-0.46) | -0.07 (-0.33,0.19) |
| n-6 fatty acids | -1.17 (-1.28,-1.06) | -0.04 (-0.27,0.20) | -1.43 (-1.55,-1.31) | -0.02 (-0.29,0.25) |
| PUFA | -1.14 (-1.25,-1.03) | -0.05 (-0.28,0.19) | -1.38 (-1.50,-1.26) | -0.02 (-0.29,0.25) |
| Saturated fatty acids | -1.34 (-1.44,-1.24) | 0.01 (-0.21,0.22) | -1.49 (-1.60,-1.38) | -0.02 (-0.26,0.23) |
| Total fatty acids | -1.08 (-1.19,-0.98) | -0.01 (-0.24,0.22) | -1.23 (-1.34,-1.11) | -0.02 (-0.28,0.24) |
| **Other metabolites** |  |  |  |  |
| Acetate | -0.56 (-0.65,-0.47) | -0.05 (-0.24,0.13) | -0.48 (-0.57,-0.39) | 0.11 (-0.08,0.30) |
| Acetoacetate | -0.43 (-0.49,-0.36) | -0.19 (-0.33,-0.04) | -0.36 (-0.44,-0.29) | -0.22 (-0.37,-0.07) |
| Albumin | 0.20 (0.06,0.34) | -0.32 (-0.62,-0.03) | 1.32 (1.17,1.46) | -0.36 (-0.71,-0.02) |
| Beta-hydroxybutyrate | 0.95 (0.84,1.05) | -0.28 (-0.51,-0.06) | 0.84 (0.75,0.93) | -0.26 (-0.46,-0.06) |
| Creatinine | 3.18 (3.06,3.30) | -0.08 (-0.35,0.18) | 5.40 (5.27,5.54) | 0.01 (-0.29,0.32) |
| Glucose | -0.63 (-0.73,-0.54) | -0.02 (-0.22,0.18) | -0.39 (-0.49,-0.29) | -0.08 (-0.29,0.13) |
| Glycoprotein acetyls | -0.26 (-0.37,-0.15) | 0.49 (0.24,0.73) | -0.13 (-0.25,-0.004) | 0.25 (-0.03,0.53) |

**eTable 11:** **Mean change in amino acids, fatty acids and other metabolic concentrations between 7y and 25y in females, by household social class, estimated from multilevel models**

Results shown are standardised mean change and mean difference in metabolic traits. The intercept, spline periods and coefficients for the slope index of inequality from multilevel models were used to estimate the mean change in concentration for degree level maternal education and mean difference for less than O-level. CI, confidence interval; MUFA, monounsaturated fatty acids; PUFA, polyunsaturated fatty acids; VLDL, very-low-density lipoprotein. *Conjugated linoleic acid and fatty acid chain length are modelled up to 18y only.

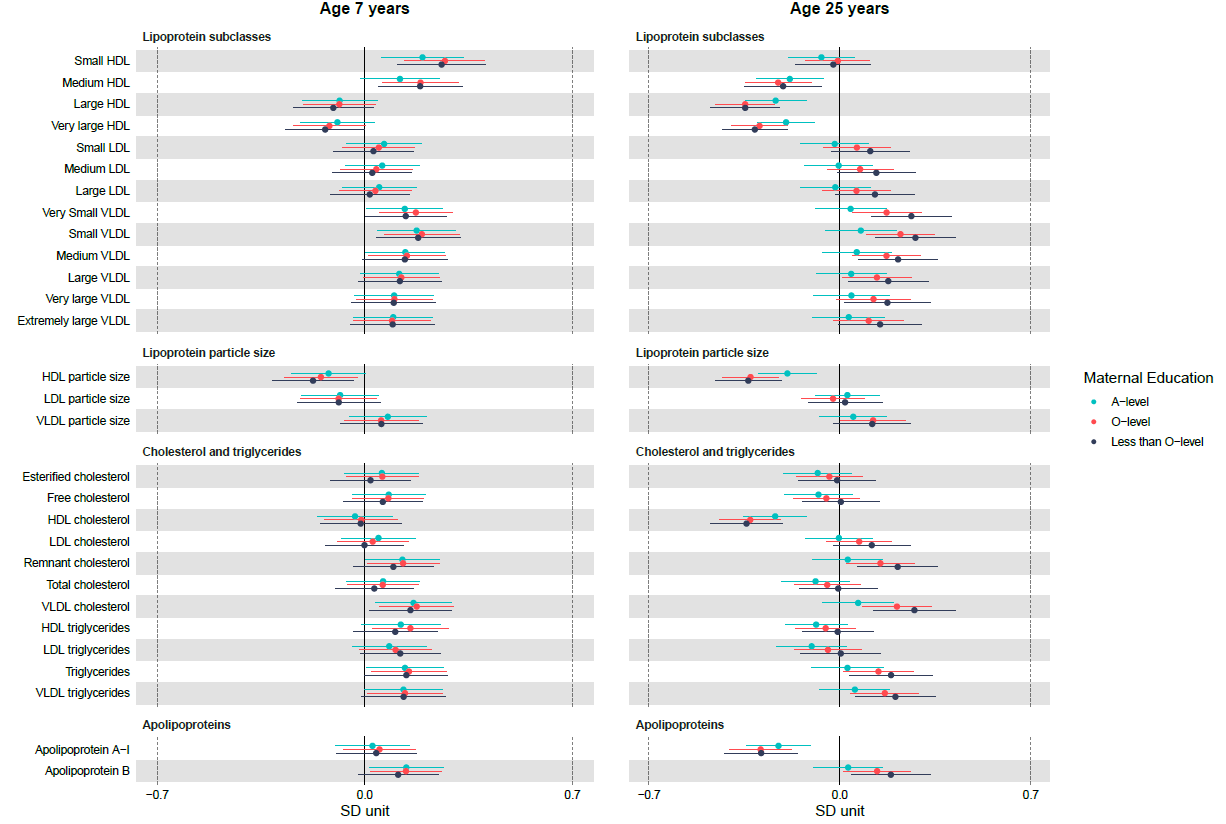

**eFigure 7: Sex-specific associations of lower maternal education categories (compared with degree level) with lipoprotein and lipid concentrations at 7y and 25y in females, estimated from multilevel models**

Results shown are standardised differences in metabolic trait comparing maternal education categories to degree level. Maternal education is categorised as less than O-level (Ordinary Level; exams taken in different subjects usually at age 15 or 16 years at the completion of legally required school attendance, equivalent to the present UK General Certificate of Secondary Education), O-level, A-level (Advanced Level; exams taken in different subjects usually at age 18 years), or university degree or above. HDL, high-density lipoprotein; LDL, low-density lipoprotein; SD, standard deviation; VLDL, very-low-density lipoprotein.

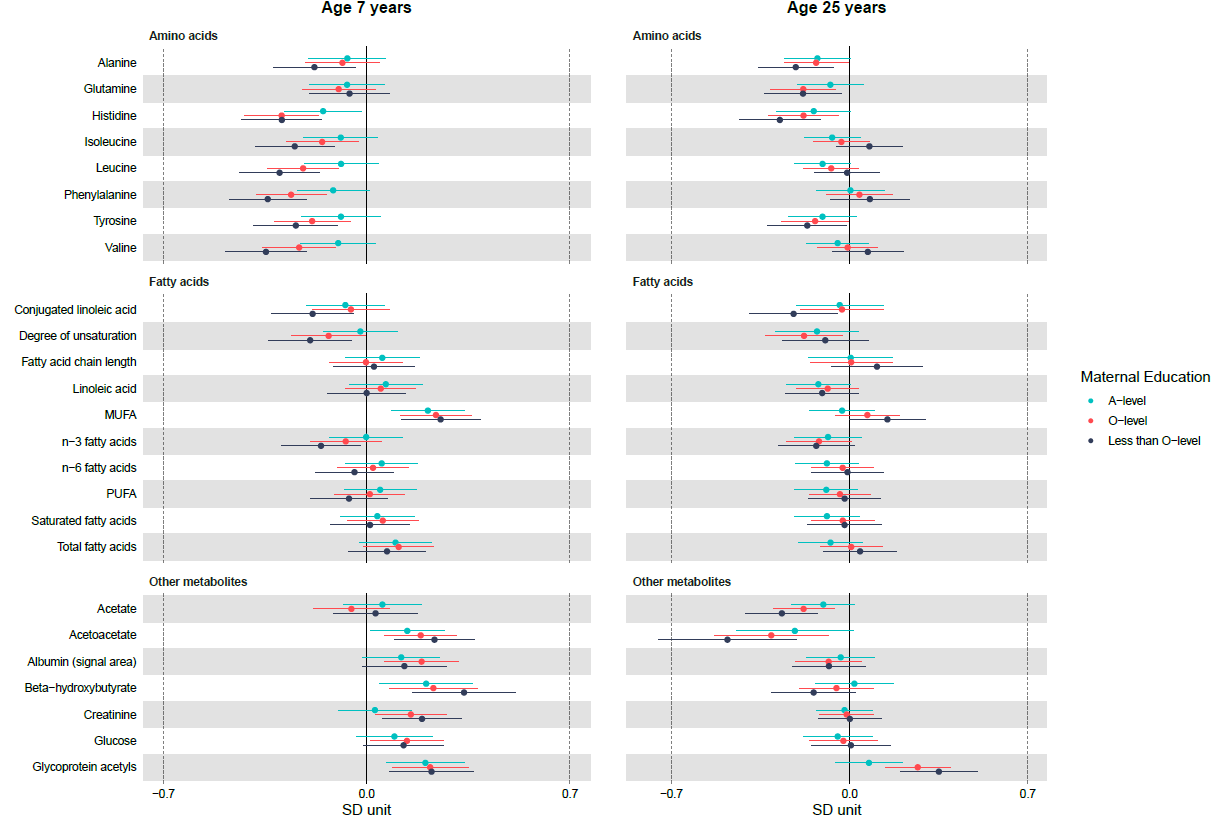

**eFigure 8: Sex-specific associations of lower maternal education categories (compared with degree level) with amino acids, fatty acids and other metabolic concentrations at 7y and 25y in females, estimated from multilevel models**

Results shown are standardised differences in metabolic trait comparing maternal education categories to degree level. Maternal education is categorised as less than O-level (Ordinary Level; exams taken in different subjects usually at age 15 or 16 years at the completion of legally required school attendance, equivalent to the present UK General Certificate of Secondary Education), O-level, A-level (Advanced Level; exams taken in different subjects usually at age 18 years), or university degree or above. MUFA, monounsaturated fatty acids; PUFA, polyunsaturated fatty acids; SD, standard deviation; VLDL, very-low-density lipoprotein. *Conjugated linoleic acid and fatty acid chain length are modelled up to 18y only.

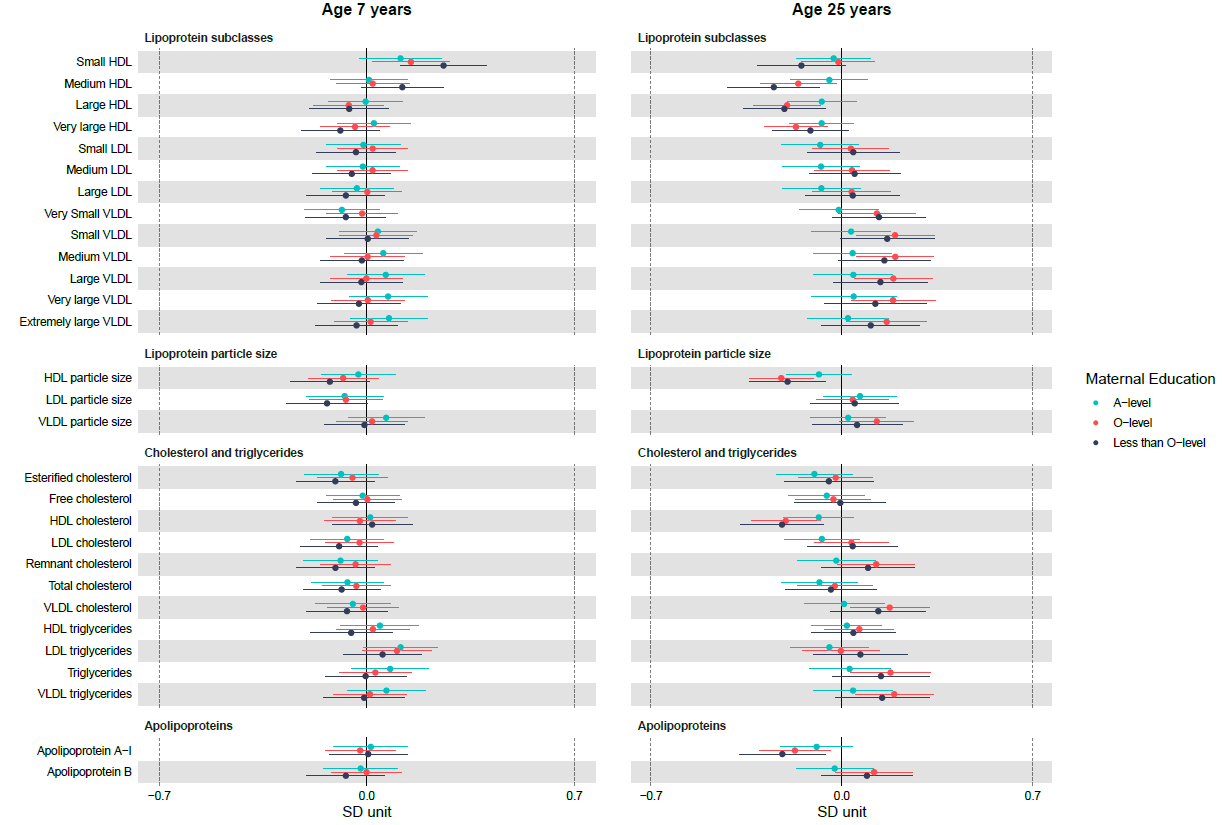

**eFigure 9: Sex-specific associations of lower maternal education categories (compared with degree level) with lipoprotein and lipid concentrations at 7y and 25y in males, estimated from multilevel models**

Results shown are standardised differences in metabolic trait comparing maternal education categories to degree level. Maternal education is categorised as less than O-level (Ordinary Level; exams taken in different subjects usually at age 15 or 16 years at the completion of legally required school attendance, equivalent to the present UK General Certificate of Secondary Education), O-level, A-level (Advanced Level; exams taken in different subjects usually at age 18 years), or university degree or above. HDL, high-density lipoprotein; LDL, low-density lipoprotein; SD, standard deviation; VLDL, very-low-density lipoprotein.

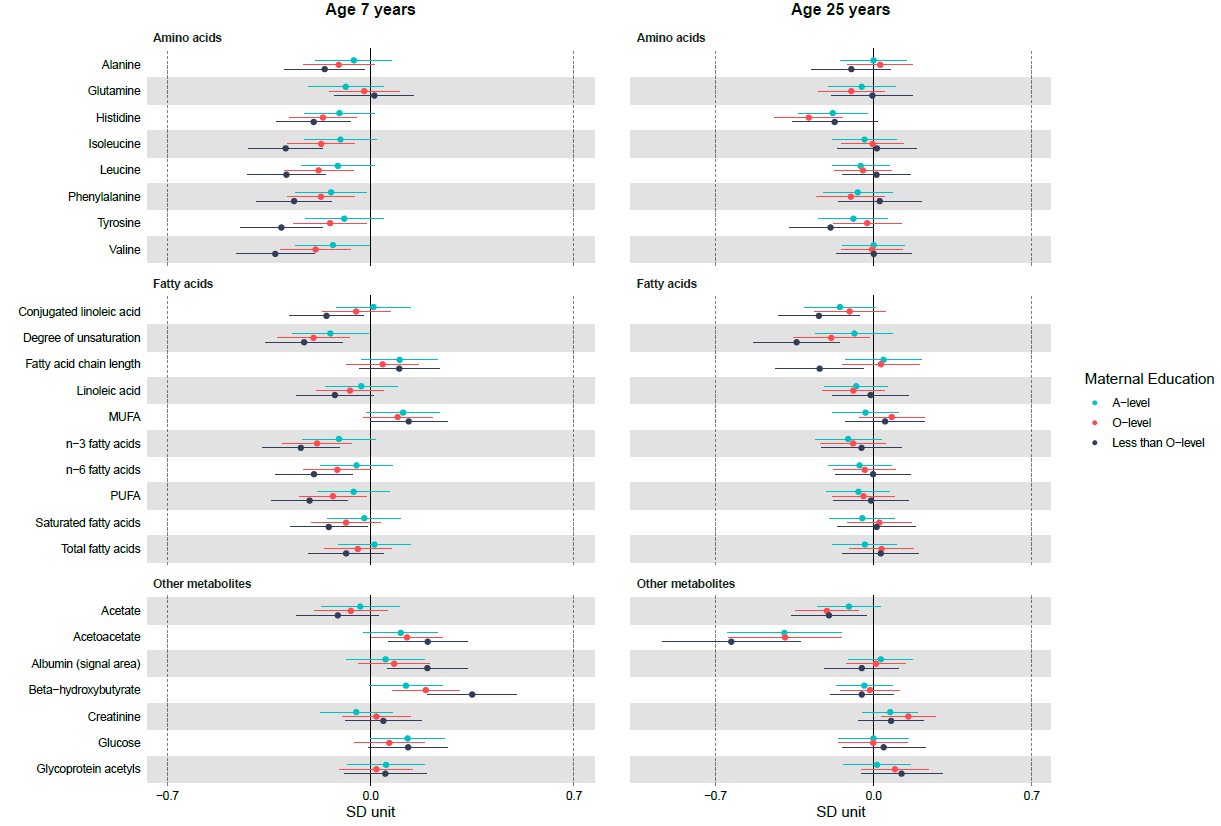

**eFigure 10: Sex-specific associations of lower maternal education categories (compared with degree level) with amino acids, fatty acids and other metabolic concentrations at 7y and 25y in males, estimated from multilevel models**

Results shown are standardised differences in metabolic trait comparing maternal education categories to degree level. Maternal education is categorised as less than O-level (Ordinary Level; exams taken in different subjects usually at age 15 or 16 years at the completion of legally required school attendance, equivalent to the present UK General Certificate of Secondary Education), O-level, A-level (Advanced Level; exams taken in different subjects usually at age 18 years), or university degree or above. MUFA, monounsaturated fatty acids; PUFA, polyunsaturated fatty acids; SD, standard deviation; VLDL, very-low-density lipoprotein. *Conjugated linoleic acid and fatty acid chain length are modelled up to 18y only.

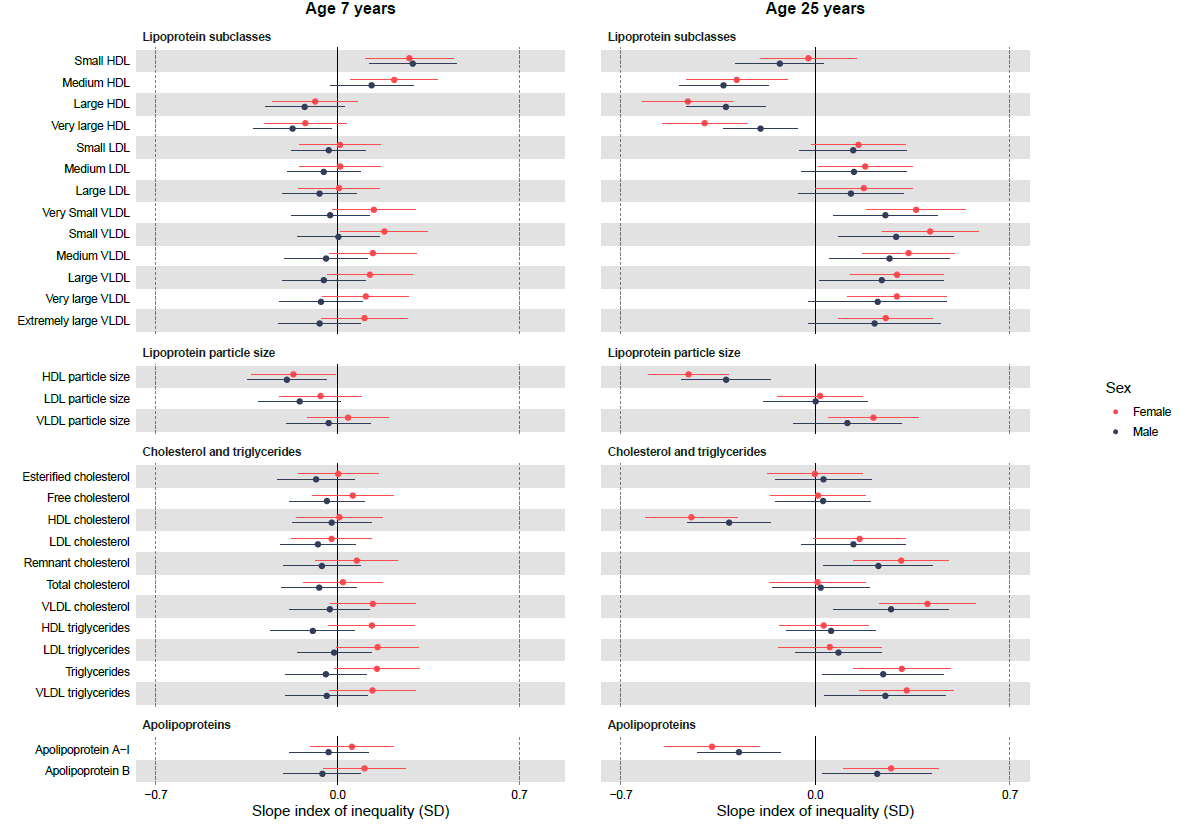

**eFigure 11: Weighted sex-specific associations of maternal education with lipoprotein and lipid concentrations at 7y and 25y, estimated from multilevel models**

Slope index of inequality represents the mean difference in SDs of the outcome between the individuals of lowest and highest socioeconomic position on the hypothetical underlying continuous distribution of paternal education. Results shown are standardised mean differences in metabolic traits comparing lowest paternal education (less than O-level) with highest paternal education (degree level) with whiskers representing 95% confidence intervals. HDL, high-density lipoprotein; LDL, low-density lipoprotein; SD, standard deviation; VLDL, very-low-density lipoprotein.

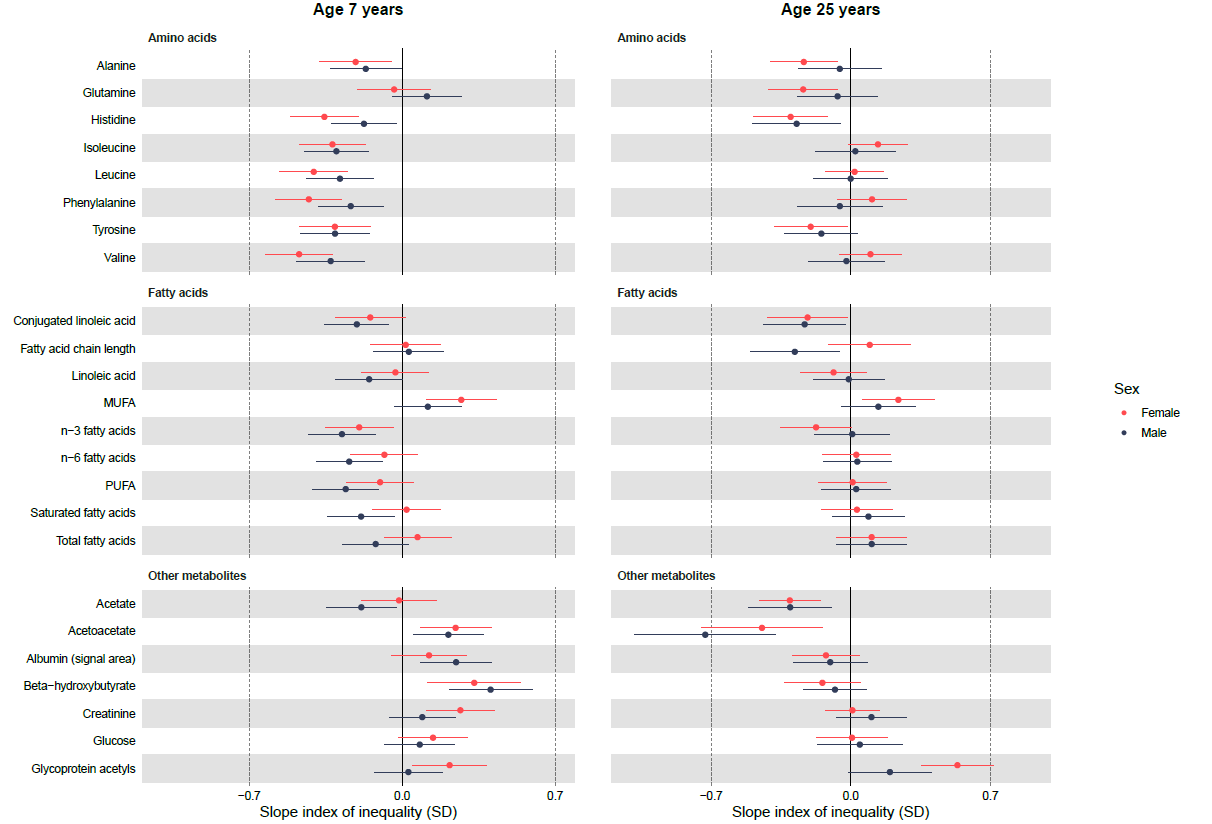

**eFigure 12: Weighted sex-specific associations of maternal education with amino acids, fatty acids and other metabolic concentrations at 7y and 25y, estimated from multilevel models**

Slope index of inequality represents the mean difference in SDs of the outcome between the individuals of lowest and highest socioeconomic position on the hypothetical underlying continuous distribution of maternal education. Results shown are standardised mean differences in metabolic traits comparing lowest paternal education (less than O-level) with highest paternal education (degree level) with whiskers representing 95% confidence intervals. MUFA, monounsaturated fatty acids; PUFA, polyunsaturated fatty acids; SD, standard deviation; VLDL, very-low-density lipoprotein. *Conjugated linoleic acid and fatty acid chain length are modelled up to 18y only.
